# Longitudinal analysis within one hospital in sub-Saharan Africa over 20 years reveals repeated replacements of dominant clones of *Klebsiella pneumoniae* and stresses the importance to include temporal patterns for vaccine design considerations

**DOI:** 10.1101/2023.09.26.23296137

**Authors:** Eva Heinz, Oliver Pearse, Allan Zuza, Sithembile Bilima, Chisomo Msefula, Patrick Musicha, Patriciah Siyabu, Edith Tewesa, Fabrice E Graf, Rebecca Lester, Samantha Lissauer, Jennifer Cornick, Joseph M Lewis, Kondwani Kawaza, Nicholas R Thomson, Nicholas A Feasey

## Abstract

Infections caused by multidrug-resistant gram-negative bacteria present a severe threat to global public health. The WHO defines drug-resistant *Klebsiella pneumoniae* as a priority pathogen for which alternative treatments are needed given the limited treatment options and the rapid acquisition of novel resistance mechanisms by this species. Longitudinal descriptions of genomic epidemiology of *Klebsiella pneumoniae* can inform management strategies but data from sub-Saharan Africa are lacking.

We present a longitudinal analysis of all invasive *K. pneumoniae* isolates from a single hospital in Blantyre, Malawi, southern Africa, from 1998-2020, combining clinical data with genome sequence analysis of the isolates. We show that after a dramatic increase in the number of infections from 2016 *K. pneumoniae* becomes hyperendemic, driven by an increase in neonatal infections. Genomic data show repeated waves of clonal expansion of different, often ward-restricted, lineages, suggestive of hospital associated transmission. We describe temporal trends in resistance and surface antigens, of relevance for vaccine development.

Our data highlight a clear need for new interventions to prevent rather than treat *K. pneumoniae* infections in our setting. Whilst one option may be a vaccine, the majority of cases could be avoided by an increased focus on and investment in infection prevention and control measures, which would reduce all healthcare associated infections and not just one.

## Background

The burden of deaths in children under the age of five, including those due to neonatal sepsis, is especially high in low-and middle-income countries (LMICs)(1); and one of the main pathogens causing these is *Klebsiella pneumoniae* (*Kpn*)(2). Until the last decade, *Kpn* was typically readily treatable with beta-lactam antimicrobials, in particular third generation cephalosporins (3GC). The effectiveness of 3GC is lost when these bacteria acquire extended-spectrum beta-lactamase enzymes (ESBLs). For *Kpn* there has been a drastic increase in 3GC-resistance by acquisition of ESBL enzyme-encoding plasmids; the proportion of *Kpn* hospital isolates that are ESBL-producing increased from 12% to >90% in parts of sub-Saharan Africa (sSA) in the last decade(3). The main remaining treatment option are carbapenems; these however are uncommonly available in sSA, rendering *Kpn* effectively untreatable in large parts of Africa. This is particularly problematic for neonatal sepsis caused by *Kpn*, and there is increasing evidence from large studies such as BARNARDS(2) and CHAMPS(4) that the burden of disease caused by *Kpn* falls most heavily on this patient group. Treatment options for neonates are even more limited than for other age groups as therapy needs to be intravenous and some antimicrobial classes are contraindicated, however delay in establishing appropriate therapy is even more dangerous.

The population structure of *Kpn*, similar to other opportunistic pathogens that thrive in the environment, is highly complex(5). Understanding long-term population trends of clinically relevant lineages at both a local and global scale, and which antimicrobial resistance plasmids and elements they encode, is potentially relevant to guiding treatment, infection prevention strategies and vaccine development. However, few longitudinal studies of *Kpn* population have been published so far, and the majority have come from collections established in high-income settings or South East Asia(6–10), whilst data from Africa and South America remain sparse. Low-income countries in sub-Saharan Africa are particularly underrepresented in ‘global’ collections of pathogen genome data, despite often carrying the highest burden of disease(2).

Whilst *Kpn* has a global distribution, there are clear geographical concentrations of dominant lineages and resistance genes, likely reflecting local antimicrobial pressure, patient populations, and a range of other determinants. Developing a more representative picture of the global epidemiology of *Kpn* is relevant to the development of vaccines targeting *Kpn*, to ensure that the lineages and associated LPS O-antigens and capsular polysaccharide antigens most prevalent in countries with the highest burden of disease are targeted. A better understanding of the genomic epidemiology of *Kpn* in low-income countries is also crucial to advocate for access to antimicrobials, in particular newer agents, which is often profoundly limited by cost, leaving isolates that can be readily treated in high-or middle-income countries *de facto* untreatable. Further, knowing at a facility level where problematic hotspots of transmission are within a hospital setting can help focus infection prevention and control (IPC) efforts.

To address this urgent need for a better understanding of the *Kpn* population in sSA, we performed whole-genome sequencing on all bloodstream isolates phenotypically identified as *Kpn* from Queen Elizabeth Central Hospital (QECH) in Blantyre, Malawi, which has undertaken sentinel surveillance of bacteraemia and meningitis in partnership with the Malawi-Liverpool Wellcome Programme (MLW) since 1998(3). Our analyses include all isolates, irrespective of resistance profile or patient demographic, from 1996 to 2020, with no change in sampling strategy from 2000 - 2020. We report the shifts in resistance profile and population structure over time and highlight the relevance to consider trends over time and not just total numbers for considerations of new drug regiments or vaccine target selection.

## Results

### The QECH bloodstream isolate collection

QECH is the main referral hospital for the southern region of Malawi and the main hospital for Blantyre, the second largest city of Malawi (population 800,024 at the 2018 census). It has approximately 1,000 beds and receives around 10,000 adult medical and 30,000 paediatric medical admissions per year. A quality assured diagnostic microbiology service was initially implemented in partnership with MLW at QECH in 1998. Laboratory blood culture was initially manual, and an automated system was implemented in 2000 (BacT/ALERT, Biomerieux). The diagnostic microbiology service expanded to culture cerebrospinal fluid (CSF) from 2002. During the study period, all adult and paediatric medical patients with suspected sepsis or meningitis were eligible for blood and/or CSF culture at the discretion of the admitting physician, and where possible before the administration of antimicrobials. Bacterial isolates were identified by standard techniques detailed in the methods.

At the start of the period, first-line treatment against bloodstream infections was crystal-penicillin and chloramphenicol although gentamicin was used in neonates instead of chloramphenicol. This began to change with the introduction of the 3^rd^-generation cephalosporin ceftriaxone from 2004, which was rapidly adopted as a first line agent by the Department of [adult] Medicine and more slowly by the Department of Paediatrics, which continues to use penicillin and gentamicin in the management of neonatal sepsis, as per WHO guidelines(11).

Members of the order Enterobacterales including *Kpn* were identified by API biochemical index (Biomerieux, France). *Klebsiella pneumoniae* is known to comprise a species complex including *Klebsiella pneumoniae subsp. pneumoniae* as well as several closely related species like *K. variicola* and *K. quasipneumoniae* and their respective subspecies(12). These can only be identified to species complex level via API, and we therefore refer to all members of the *K. pneumoniae* species complex as *Kpn* and specify when referring to specific (sub)species (Fig 1A).

**Fig 1.**
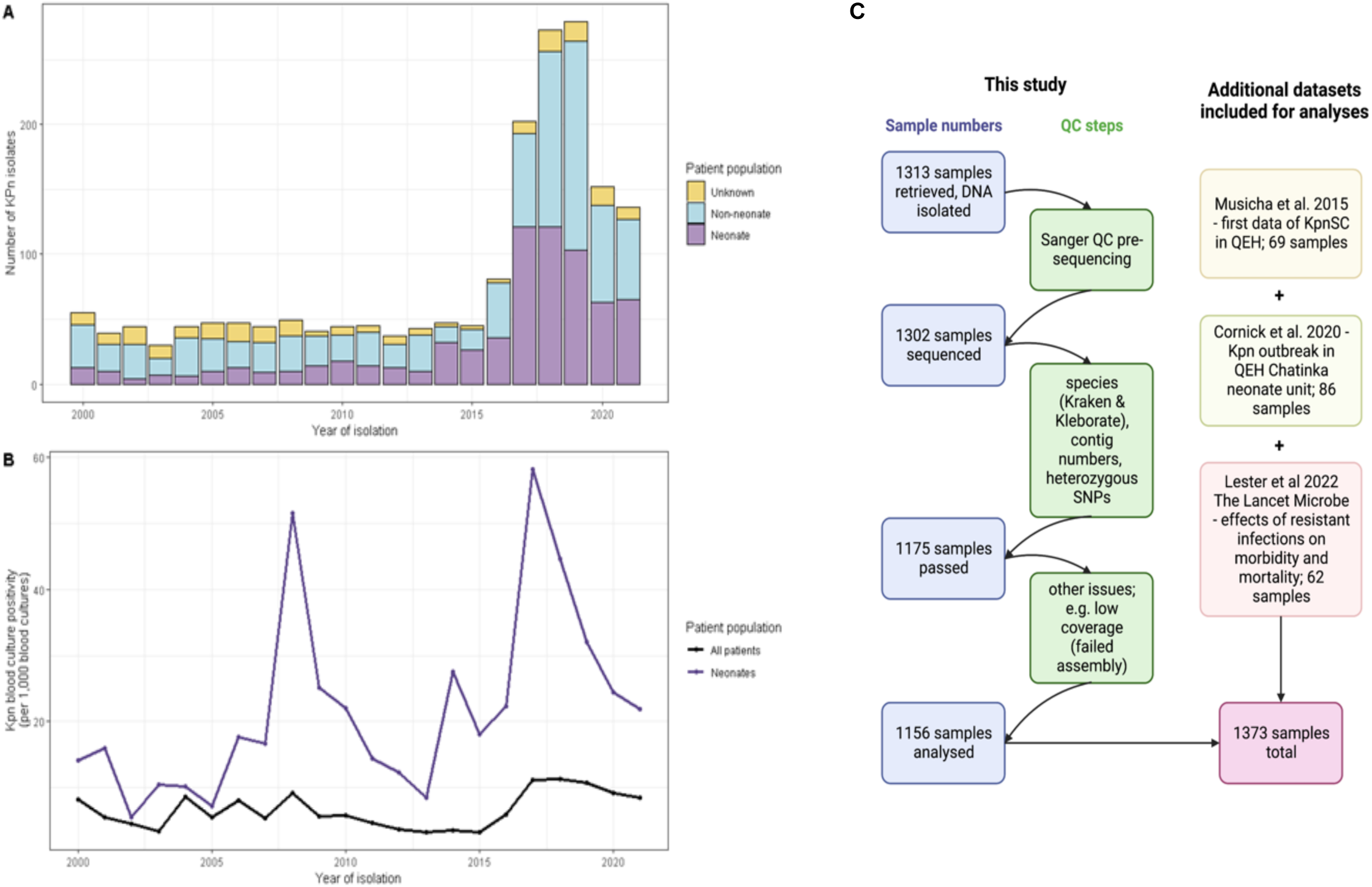
*Kpn* cases at QECH. (**A**) Numbers of *Kpn* cases per year at QECH. Bars represent the crude frequency of *Kpn* infection for each year from 1998 - 2021, with the different colours representing the different age groups of the patients. (**B**) Blood culture positivity rate (per 1,000 blood cultures) of *Kpn* in neonates and the entire patient population.

### Dramatic increase in Kpn cases from 2016

The crude frequency of *Kpn* cases was high at the start of the period with 121 cases in 1998, although this overlapped with a year of active surveillance of febrile adult patients(13) and we thus focused longitudinal analyses from 2000 onwards. The number of cases from 2000 - 2015 was stable with a mean of 44 cases per year (Fig 1A). This increased to a mean of 187 cases per year from 2016-2021 with a peak of 279 in 2019.

Neonatal infection with *Kpn* accounted for 39.3% of all *Kpn* infections, even though only 11.2% of all blood cultures were taken from this patient group. The overall rate of *Kpn* isolation was 24.9 per 1000 blood cultures for neonates, 4.3 per 1000 blood cultures for other patients (rate ratio of neonates vs non-neonates 5.11 [95% CI 4.66 - 5.61]). The increase in *Kpn* cases post 2016 was driven by an increase in neonatal infections (Fig S1A-1G); there were peaks in the *Kpn* blood culture isolation rate in neonates in 2008, 2014 and 2017 which suggest outbreaks or ongoing transmission on the neonatal unit (Fig 1B), and our genomic analyses further support hospital transmission given the close genetic relationships of the respective isolates. Though the blood culture positivity rate was increased for most age groups in the period from 2016-2021, for neonates this increase was particularly marked (33.5 per 1000 blood cultures in the period 2016-2021 compared to 15.4 per 1000 blood cultures in the period 1998-2015; Fig S1A-1D). As well as the peak in neonatal cases, there was also a much smaller peak in the adult age group, which shifted from the 30-35 year age group in the period from 1998- 2015 to the 35-40 year age group in the period from 2016-2021 (Fig S1E-1H).

### Increase in *Kpn* numbers driven by temporally defined expansions of sequence types (STs)

We retrieved 1313 isolates not previously sequenced from the QECH *Kpn* collection, 1302 of which could be recovered for growth and yielded enough DNA for sequencing; and 1156 of these passed all quality control steps for further analysis (details see methods; Fig S2). In addition, we included 217 samples from previously published studies of this collection(14–16), resulting in 1373 sequenced genomes analysed in this study (Fig 1C).

The majority (1274/1373, 92.8%) belong to *Klebsiella pneumoniae* subsp. *pneumoniae* with only very small proportions representing other (sub)species, including *K. quasipnemoniae* subsp. *quasipneumoniae* (10/1373, 0.7%), *K. quasipneumoniae* subsp. *simillipneumoniae* (46/1373, 3.4%) and *K. variicola* subsp. *variicola* (43/1373, 3.1%). Based on phenotypic microbiology data (Fig 1A and 1B) we observed a drastic increase in isolates from neonatal infection in 2016/2017 which is reflected in our sequenced collection.

Assessing diversity of STs over time highlights that the increase in *Kpn* infections represents repeated increase and subsequent decrease in numbers of different multilocus sequence types (Fig 2A). Whilst the earlier years show a diverse composition of STs with small numbers of each (n < 15 total), the increase in total *Kpn* numbers is mainly driven by one or two major STs, with these peaks spanning approximately 1.5 to 2 years each. We furthermore observe that after the increase of isolates of the major STs, each of them reverts to their original very low numbers (Fig 2A).

**Fig 2.**
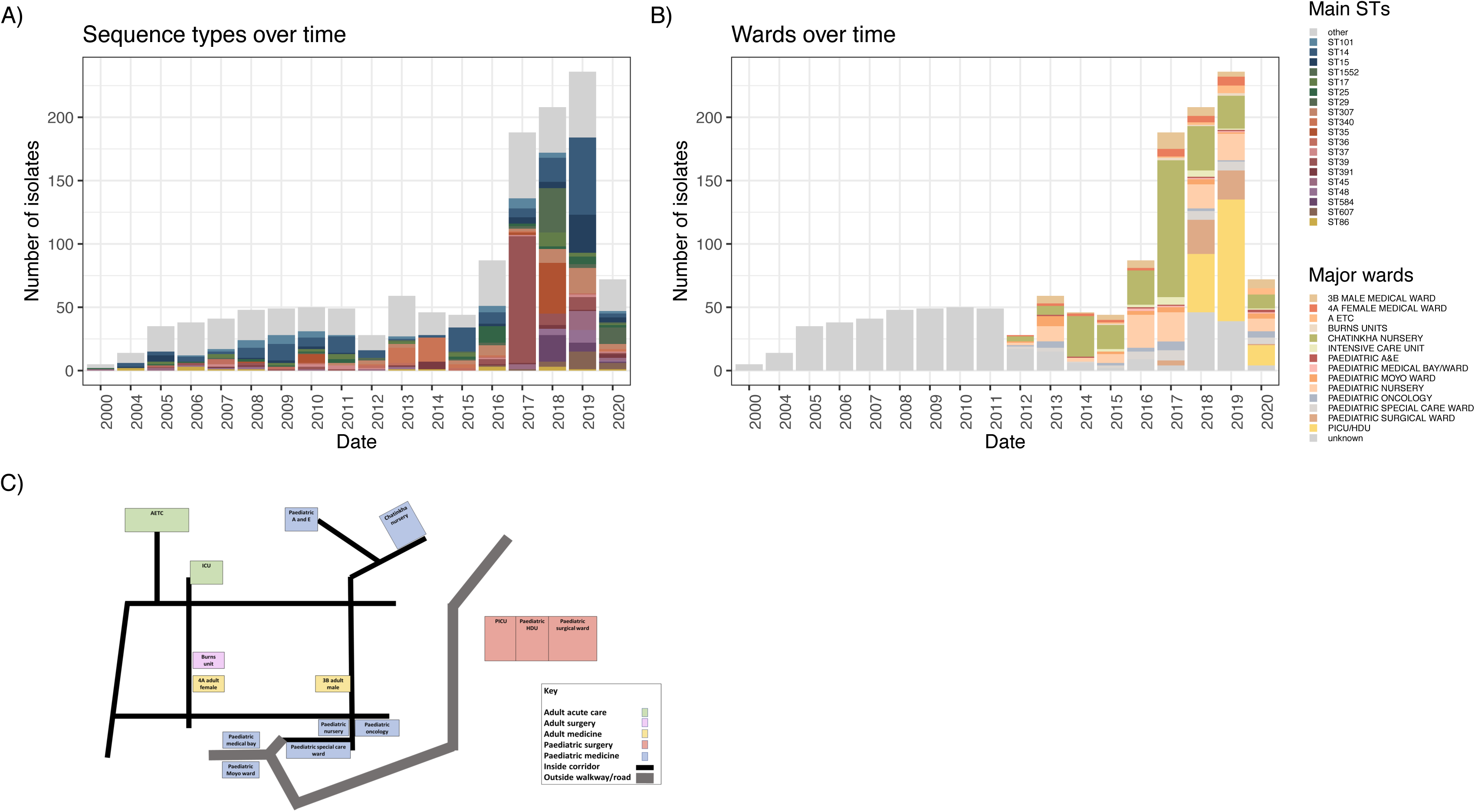
Uneven distribution of STs and cases at QECH wards indicate hospital-acquired infections. (**A**) Main STs and (**B**) wards the samples were derived from over time. (**C**) A schematic map of QECH with the wards samples of our study were derived from highlighted. AETC – Adult emergency and Trauma Centre; Paediatric A&E – paediatric accident and emergency; PICU – paediatric intensive care unit; HDU – high dependency unit.

A previous study(15) highlighted an outbreak of ST340 across the neonatal unit which is reflected here, where it comprises 31 out of 105 isolates in 2013 & 2014 (29.5%; Fig 2A). We then see an increase of the numbers of ST14 (19 / 44 in 2015; 43.2%) which is present at a lower proportion before and after this period; followed by an increase in ST25 (13 / 87 in 2016; 14.9%) which again is present at lower proportions before and after. The year 2017 then saw a steep increase in *Kpn* infections overall; mainly driven by an expansion of ST39 which comprises more than half the samples at the time (100/188 in 2017; 53.2%). This clone however again retreats back to low numbers, with a few lineages dominating, in particular ST1552 (35 / 208 in 2018; 16.8%) and ST35 (40 / 208 in 2018; 19.2%). The year 2019 then sees another increase in ST14 (61 / 236 in 2019; 25.8%) as well as ST15 (30 / 236 in 2019; 12.7%), which is a known high-risk clone predominantly found in South East Asian collections where it has been linked to an early spread of carbapenem resistances via *bla*_NDM_ genes(17,18).

### Isolation of *Kpn* from neonatal and paediatric wards account for observed changes, whilst adult infections remain stable over time

The patterns over time differ significantly between wards (Fig 2B), suggesting the increase in *Kpn* infections could represent a pattern of repeated ward-specific outbreaks, rather than a generalised increase across all wards (Fig 2B). This pattern of ward-focused increases in isolate numbers was repeatedly observed on the Chatinkha neonatal unit and the surgical PICU/HDU, where most of the changes in numbers of isolates occur. An indication of local outbreaks is further supported by analysing the major STs, which increase at defined times in specific wards (Fig 2B and 2C; Fig S3). To investigate the impact of closely related isolates (<5 core SNP distance) that are likely within-hospital transmissions on the number of infections overall, we analysed a core genome tree for each of the three species and identified closest-neighbour SNP distances in a root-to-tip approach(19) (Fig 3). This highlighted that most highly clonal lineages (defined as <5 SNPs) are derived from *K. pneumoniae* subsp. *pneumoniae* (Fig 3A) representing 402 of 1165 isolates as a conservative estimate based on whole-species analyses, with 43% of the *K. quasipneumoniae* subsp. *similipneumoniae* (20 of the 46 isolates in 6 lineages, Fig 3B) and 25% of the *K. variicola* isolates (10 of the 40 isolates of this species, Fig 3C); and no <5 SNP related isolates of *K. quasipneumoniae* subsp. *quasipneumoniae*.

**Fig 3.**
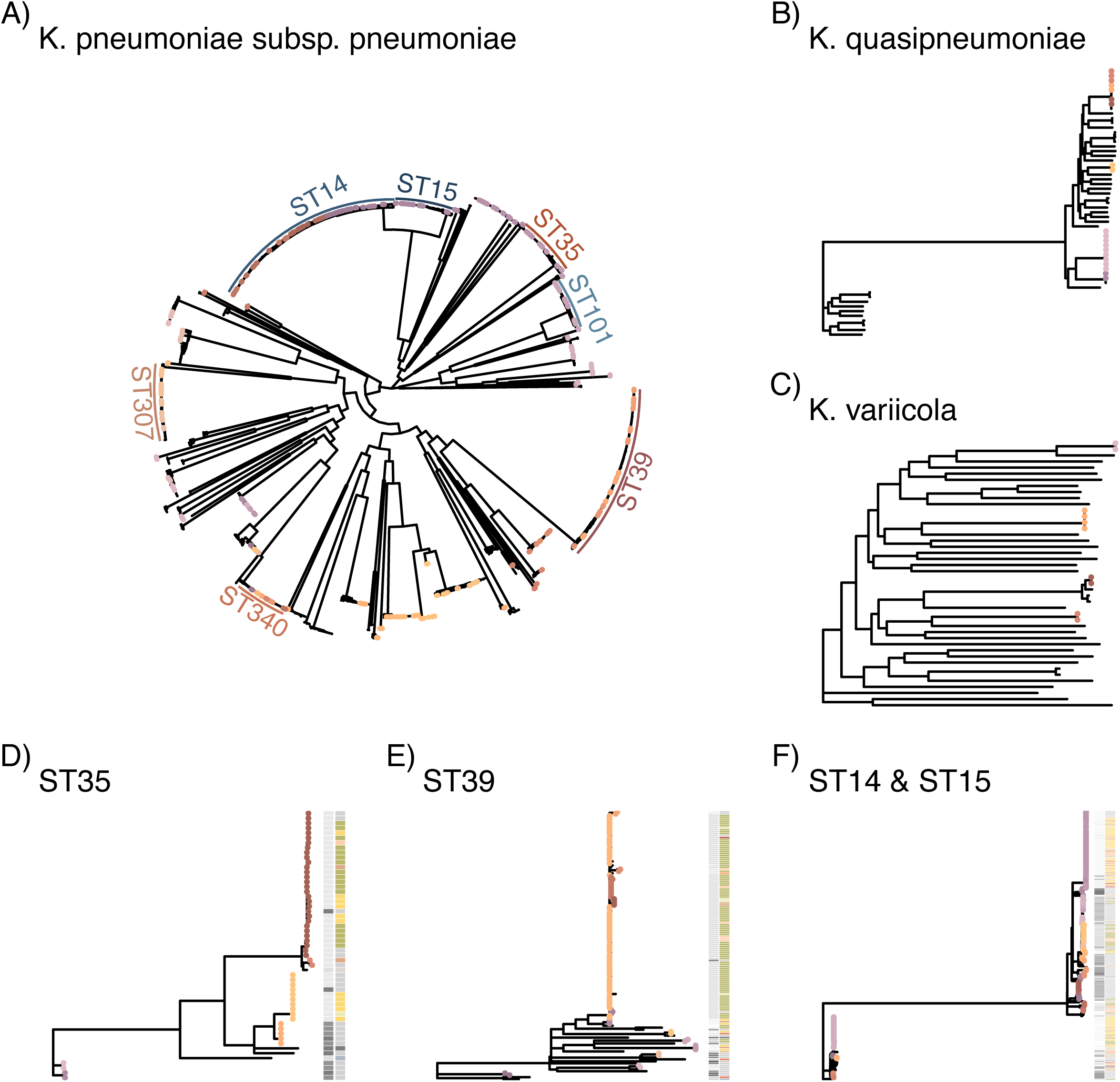
Most cases suspected to be caused by within-ward transmissions. Mapping trees against reference genomes for (**A**) *K. pneumoniae* subsp. *pneumoniae* excluding STs represented with only a single isolate in the dataset, (**B**) *K. quasipneumoniae* and (**C**) *K. variicola*. (**D-E**) show ST-specific mapping of the relevant isolates against references for ST35 (**D**), ST39 (**E**), ST14/ST15 (**F**). The coloured tips indicates isolates with <5 SNPs distance to their nearest neighbour based on a root-to-tip approach to identify related clusters of isolates(19). The colour ring in (**A**) shows the main STs, the columns in **D-F** show year of isolate (left column) and major ward (right column).

Despite removing the single-isolate STs for this analysis, our dataset still contains a high number of *K. pneumoniae* subsp. *pneumoniae* STs, which results in a highly diverse dataset. As this diversity has potential to impact on tree resolution depending on reference used(20), we investigated three key STs in more detail. We focused on ST35, ST39, and the closely related ST14 and 15 together, whose pattern of accumulation in a ward and year strongly indicates multiple hospital-associated transmissions (Fig S3), in more detail by mapping against strain-specific reference genomes (Fig 3A). ST39 and ST35 are two key STs when considering the per-year breakdown of isolates per ward for the neonatal unit (Chatinka; Fig S3); whilst ST14 and ST15 were identified in high numbers in the PICU/HDU in the paediatric surgical ward (Fig S3). Here, we can observe that 77% of the isolates (51/55 ST35, 110/136 ST39, 182/254 ST14-15) have their nearest neighbour within the threshold of 5 SNPs; representing 100% (17/17) of ST35 and 88.9% (80/90) of ST39 from Chatinka and 88% (57/65) ST14 and 15 from the PICU/HDU (Fig 3A). This high relatedness of isolates strongly supports within ward transmission and the argument that *Kpn* is a healthcare associated cause of neonatal sepsis. As patients are regularly transferred from Chatinkha to the PICU/HDU if surgical procedure is required, these wards are furthermore already linked by a clear transmission path via patients, and several potential between-ward transmission events are indicated in particular in ST35 and ST14/15 (Fig 3D, F).

Over the period from 2015-2018 there was an increased focus on performing blood cultures to ensure every neonate on Chatinkha nursery that met the clinical criteria for suspected neonatal sepsis was included, which may explain some of the large increase in *Kpn* cases in 2017 on the nursery. Establishing optimal IPC has been an ongoing challenge(21); specifically, handwashing has been difficult due to intermittent water supply and a lack of access to soap or hand sanitiser and there are limited numbers of cots, so neonates have often shared them. In response to the large increase in *Kpn* cases there was renewed focus on IPC; hand sanitizer and soap procurement was prioritized, deep cleans of the ward environment were performed regularly, there was increased cleaning of ward surfaces, ward traffic was limited, wooden cots are being replaced with easier to clean plastic ones, the number of neonates sharing a cot was reduced and oxygen delivery devices are being replaced with single use items.

### Antimicrobial resistance and implications for treatment

Phenotypic resistance profiles of the strains, where available, mirrors the very concerning trends reported previously for *Kpn* in QECH(3) (Fig S4, Table S4). Resistance against the frontline treatments is widespread; a particular concern is a high proportion of isolates with ceftriaxone, gentamicin and cotrimoxazole resistance from 2012 onwards with very little change over time (Fig S4A-C; >75% of tested isolates were resistant to these agents each year 2012 – 2020). We notice some variation in co-amoxiclav (augmentin: amoxicillin + clavulanic acid) susceptibility profiles but from 2019 again show >75% of tested isolates as resistant (Fig S4A). Importantly, we notice that a trend of increased numbers of chloramphenicol susceptible isolates (Fig 4B, Fig S4G), which has previously reported in Malawi(3), is also explicit in our *Kpn* collection in particular in recent isolates (38.7%, 51.5%, 72.2% isolates sensitive for 2018-2020). Presence of *catA1* or *catA2* agree well with phenotypic resistance whereas presence of *catB4* did not as shown in Fig 4B; where we see a large number of isolates with sensitive phenotype, no *catA1/2* but *catB4*, whereas only a small fraction of isolates has resistant phenotype, no *catA1/2* but *catB4* where we hypothesize a different mechanism is likely driving the resistant phenotype. For isolates with resistant phenotype and encoding both *catA1/2* and *catB4* the resistance is presumed to be driven by *catA1/2*. This agrees with *catB4* representing a non-functional truncated variant of *catB3* as was recently reported. We furthermore note a sensitivity of 50% or higher in the majority of years measured in ciprofloxacin (6/9 years from 2012 to 2020, including 2017, 2018 and 2020; Fig S4E, Table S4), which together with chloramphenicol thus represent two antimicrobials that might thus be valid treatment options.

**Fig 4.**
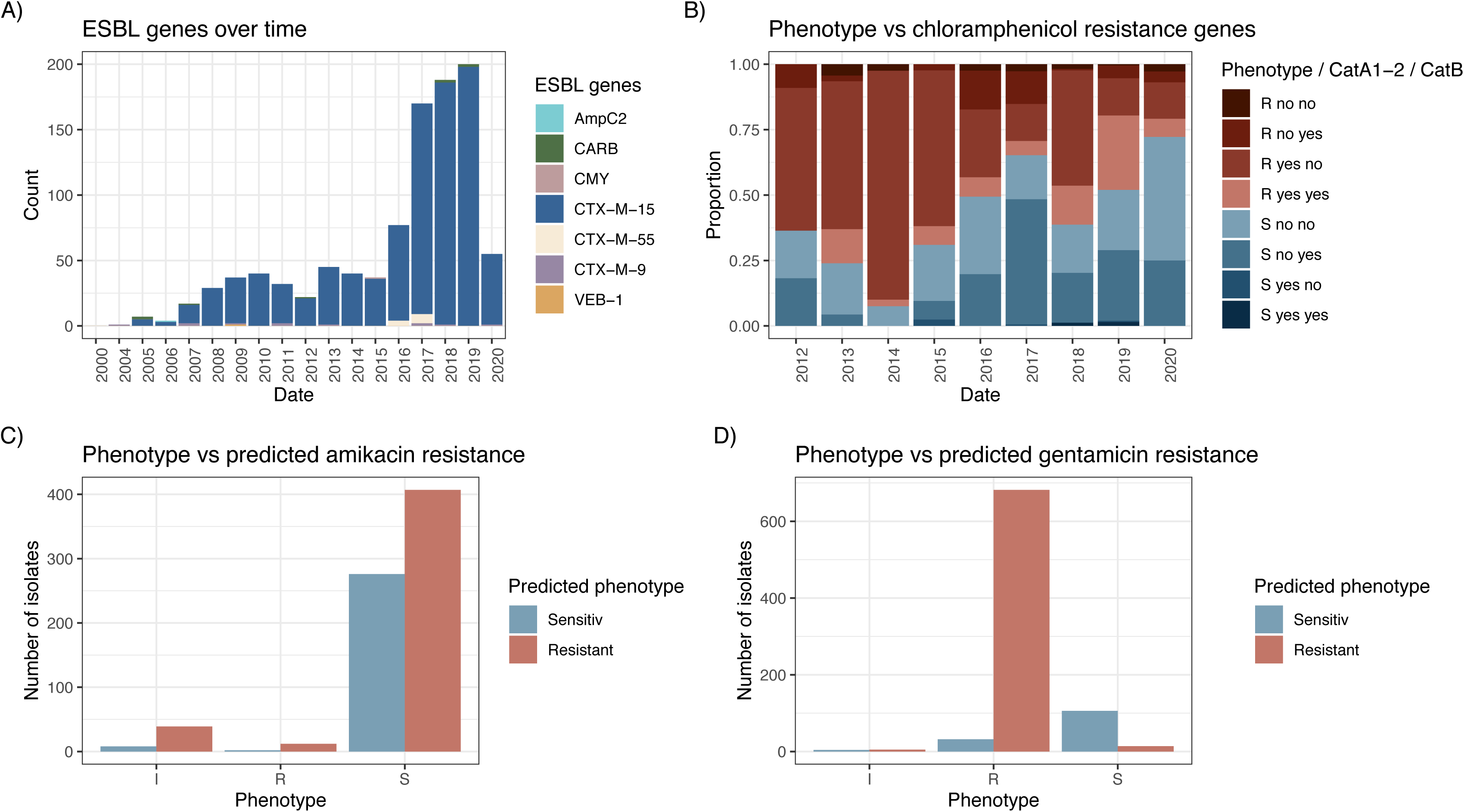
Resistance mechanisms only explain beta-lactam, not aminoglycoside, phenotypes. (**A**) The distribution of ESBL genes over time illustrating the dominance of *bla_CTX-M-15_*. (**B**) Phenotypic chloramphenicol resistance profiles proportional over time and the presence or absence of *catA1* and/or *catA2* compared to *catB4*, the recently re-characterised non-functional CAT enzyme. (**C**) and (**D**) Predicted aminoglycoside resistance was based on the presence of an Aac6’-Ib-cr or Aac6’-Ib gene (AAC(6’)-I) for amikacin and Aac3_IIa (AAC(3)-II) or AadB (ANT(2”)-I) for gentamicin. The x-axis shows the samples based on phenotypic resistance profiles (intermediate, sensitive, resistant) against (**C**) amikacin and (**D**) gentamicin, and the bars show number of isolates in the respective category that would have been predicted as sensitive (blue) or resistant (red) based on the genes as above.

Resistance to 3GC was widespread (83% resistance to cefpodoxime/ceftriaxone by 2020; Fig S4A) and almost exclusively driven by acquisition of the *bla_CTX-M-15_* gene, which continuously outcompetes other ESBL-encoding genes that are occasionally present in the *Kpn* population, but which are only sporadically detected (Fig 4A, Table S2). The number of plasmid replicons per isolate stabilised after the first increase in numbers around 2015, highlighting that a few large AMR plasmids are likely responsible for the majority of AMR-isolates (Fig S5C, Fig S6A, Table S3). Strains with a moderate number of plasmid replicons dominate the population (0- 5) and this remains stable over time, with only a small number of isolates with more than five plasmid replicons associated that remains at similar numbers over time (Fig S6A). This indicates that the change to a predominantly ESBL^+^ population was the key event, with the population then remaining stable over time with respect to plasmid diversity and numbers of resistance genes (Fig S6B).

Carbapenem resistance genes are still only rarely observed; we identified two samples encoding for *bla_NDM-1_*; one in a ST39 in 2016 and one in a ST45 in 2019, perhaps unsurprisingly as carbapenem usage is still comparatively unusual. We also investigated resistances against other potential alternative treatment options colistin, fosfomycin and tigecycline. Resistance to these agents are all at low levels, with only one acquired fosfomycin resistance gene detected (*fosA*) and none against colistin or tigecycline predicted in our dataset, and one isolate with predicted mutations that can confer resistance to colisin (*pmrB*), indicating that these might be viable alternatives for ESBL^+^ infections at QECH, although tigecycline is contraindicated in children under 8y (Table S2) and neither agent may not be ideal treatments for bacteraemia(22,23). Tigecycline resistance is often conferred by upregulation of transport proteins and thus challenging to predict and has also been observed during within-patient evolution(24–26); we could however not detect the described plasmid-derived antimicrobial resistance genes *tetX* in our collection(27).

Based on AMR phenotype identified by disc diffusion, amikacin remains a viable alternative to gentamicin, although there are concerns over blood brain barrier penetration in cases of meningitis and renal toxicity in settings where therapeutic drug monitoring is not possible. Phenotypic data are corroborated by a more detailed analysis of the aminoglycoside AMR genes (Fig 4C and 4D). In total 90.5% of all isolates (1243/1373) are predicted to encode one or more aminoglycoside resistance genes, however, the majority are predicted to confer resistance to streptomycin (ANT3) and gentamicin (AAC(3)-II; ANT(2”)-I; Fig 4D). Only a fraction of these strains (43.2%, 593/1373) encoded one of the genes predicted to confer resistance against amikacin (AAC(6’)-I; Fig 4C). Aminoglycoside resistance is known to be challenging to predict(28); whilst phenotypic gentamicin resistance testing matches very well with the predicted profile based on acquired genes, the presence of AAC(6’)-I does not by itself confer resistance in a large number of isolates, with only 12 isolates encoding for either of these, representing 2%, actually showed a resistant phenotype (Fig 4D; Table S4).

### Potential vaccine target stability

The *Kpn* polysaccharide antigens are of high interest as vaccine targets; in particular the large capsular antigen. As expected, we observed a very high number of different K-loci (90 predicted), as well as a diverse set of O-loci (15 predicted), the latter are known to be far less diverse in *Kpn*(29). Recent findings furthermore showed that *wbbZ,* encoded on the chromosome outside the O-locus, contributes to the diversity of the O-Ag structure displayed by *Kpn* and thus two isolates with the same O-locus can still result in different O-types based on the presence or absence of this gene. Immunologically relevant differences in O-Ag thus requires the prediction of the O-Ag locus and *wbbZ*(30), so we added this to our O-Ag type predictions (Fig S7A-D) where we thus distinguish between O1a and O1b as described by Kelly *et al*(30). Reflecting the trend observed with the ST types, we observe temporal variation in prevalence of O-types and K-types (Fig 5A and 5D, Fig S7A). The non-ESBL population again displays a much greater diversity for both types of antigen (Fig 5B and 5E, Fig S7B), whilst the ESBL population is more biased towards dominant O-and K-types (Fig 5C and 5F, Fig S7C).

**Fig 5.**
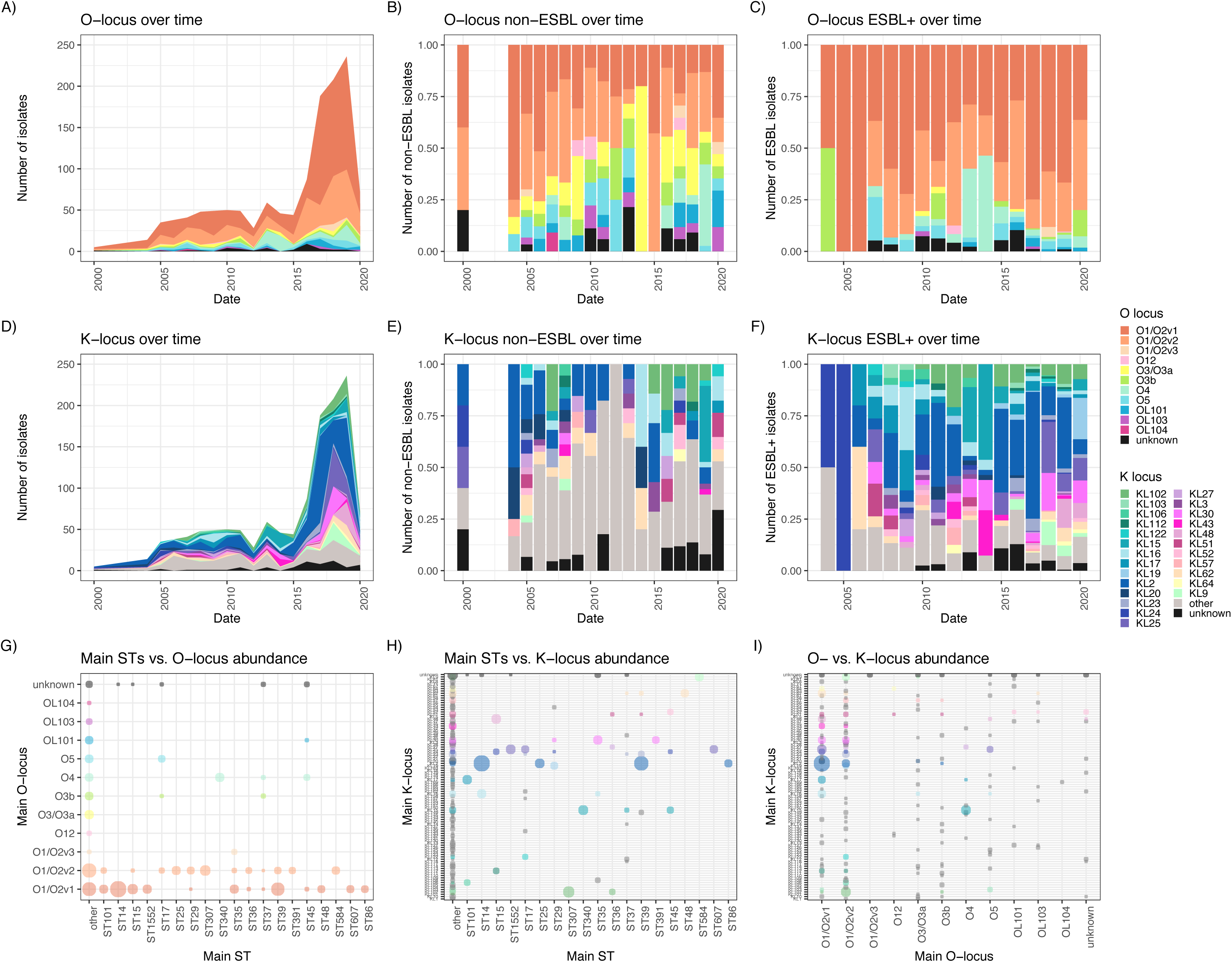
High fluctuation of putative vaccine targets over time and diversity within major STs. The distribution of O-Ag locus types (top panels) and K-locus types (lower panels), resp., over time showing total numbers (**A**) and (**D**), only isolates with ESBL resistance genes (**B**) and (**E**) or ESBL-negative isolates (**C**) and (**F**).

Beyond the dominant O1b (821/1373; 59.8%) and O2afg (184/1373; 13.4%) predicted O-antigen types, the infrequently observed type O4 comprises a substantial fraction of isolates (90/1373; 6.6%); and 35 isolates have a prediction accuracy as “Low” or “None” which can indicate either low assembly quality or novel O-Ag loci and types. In addition, we note two isolates with unknown O-Ag locus encoding for additional *wbbY* and *wbbZ* genes, indicating that more variation might be within possibly novel O-Ag loci depending on additional enzymes encoded in the chromosomal backbone (Table S5). We furthermore note 8 isolates with O1/O2 loci encoding for only *wbbZ* (not *wbbY*); whilst the recent work showed no structural differences in the O-Ag if *wbbZ* alone was added, we cannot exclude other carbohydrate enzymes on the chromosomes of our isolates that might lead to additional different structures if combined with *wbbZ* (Table S5). Of relevance, we also observe variation of O-and K-type combinations within the same sequence type (Fig 5G and H, Fig S7D) and with each other (Fig 5I, Fig S7E).

Considering hypothetical vaccine strategies and constructs, an efficacious maternally administered quadrivalent O-antigen based vaccine (O1a, O1b, O2afg, O4; including O1a as O1b-enconding isolates can switch between O1a and O1b(30)) would be expected to protect against up to 89% of cases of neonatal sepsis in Blantyre (345/388), whereas a K-antigen based vaccine would require 22 K-antigens to be represented to protect against an equal number of cases of neonatal sepsis when taking the entire timespan of our study into account (KL10, KL102, KL103, KL104, KL106, KL108, KL117, KL142, KL15, KL16, KL17, KL19, KL2, KL23, KL25, KL30, KL43, KL48, KL51, KL52, KL62, KL9); a quadrivalent K-antigen based vaccine (KL2, KL25, KL30, KL102) would cover 58% (198/388) of cases. We furthermore observe no regular cycling between a specific set of K-types, but rather that different K-types regularly emerge associated with major clones (Fig 5). Given current knowledge of >180 predicted K-types(31) and >70 determined structures so far(32–34) (likely an underestimation), and the propensity of the K-locus to recombine, it will be very challenging to predict how long a K-type vaccine would remain effective.

### Trends of resistance vs virulence determinants

Whilst most *Kpn* isolates are opportunistic pathogens with poorly understood virulence profiles, the hypermucoid phenotype is a known virulence determinant arising from hyperproduction of capsule and causing a different clinical infection type, i.e. liver abscess and meningitis in otherwise healthy individuals, often progressing very rapidly and with poor prognosis. The capsule hyperproduction is usually driven by acquisition of a virulence plasmid encoding for the capsule polysaccharide upregulation factor *rmpA2* as well as several siderophore systems (aerobactin, salmochelin, colistin) that enable iron scavenging. This phenotype is of particular concern given the recent convergence of hypervirulent with multidrug resistant isolates. Whilst we note hypervirulent isolates present (defined here as kleborate virulence score >1, i.e. encoding virulence factors in addition to the often chromosomal yersiniabactin) from the very early sampling times on in our collection (Fig S8A,B, Fig S5B), these remain sporadic; in contrast to multidrug resistant isolates (Fig S8C,D Figwhich are absent at these early sampling times but rapidly become the majority of isolates from 2007/8 onwards (Fig S8D). Whilst we note a total number of >10 isolates for both ST86 and ST268, two STs carrying the *rmpA2* gene and widely observed as hypervirulent phenotypes, these occur in different years and wards, highlighting that hospital-derived transmission is highly unlikely (Fig S8B). Whilst it is of concern that we observe some of these potentially hypervirulent isolates (encoding for *rmpA2*) also encoding an ESBL gene (22 total; 9 of these ST86, no ST268) there seems to be no selective advantage; the first isolate carrying this combination is observed in 2010 but a maximum of 4 isolates per year are observed until 2020. This small number of putative hypervirulent isolates is in accordance not only with bloodstream isolates in general but also with a study comparing carriage with infectious isolates from adults in Blantyre, where only a very small number of isolates encoding virulence determinants were identified, all of them in infectious isolates(35,36).

## Discussion

Neonatal sepsis is responsible for a high proportion of deaths in children under 5 in sSA, with *Klebsiella* by now being both the most prevalent gram-negative pathogen and increasingly hard to treat. We have analysed a longitudinal collection of routine diagnostics invasive (bloodstream and CSF) isolates of *Kpn* spanning from 1998 – 2020, during which time AMR increased drastically. We determined longitudinal patterns of patient demographics, *Kpn* population trends, vaccine targets, resistance elements and remaining treatment options. This exceptional resource, not only for sSA but also world-wide, enables us to understand the dynamics of *Kpn* over an exceptionally long time scale where the sampling was not biased towards resistance types. In particular, it allows us to see the dynamics before, during, and after the significant expansion of ESBL producing *Kpn* at maximum resolution; and highlights the need to consider not only major types based on total numbers but the temporal dynamics of vaccine targets such as cell surface polysaccharides.

*Kpn* initially showed a bimodal age distribution in patients at QECH(3) and there were concerns that increasing life expectancy would lead to a higher number of adult infections; however, this trend has yet to be seen, and adult bloodstream infections were significantly reduced overall following the widespread distribution of antiretroviral therapy(37). The stark increase in infections from 2016 onwards is almost exclusively seen in neonates and children where ward-restricted clonal expansions that are subsequently replaced with other successful clones. This suggests that context appropriate, effective, scalable infection prevention and control (IPC) targeted towards the most vulnerable patient groups in the hospital and a better understanding of *Kpn* transmission routes and reservoirs would prevent a large proportion of the cases caused by *Klebsiella* bloodstream infection. Such approaches have the potential to protect vulnerable babies against all healthcare associated infections rather than just one pathogen and are available now, unlike a vaccine strategy that will take years to develop and implement without guarantee for success.

The main sequence types in our study were ST39, ST14 and ST15. While ST39 and ST14 are not necessarily the most prevalent ones observed globally, ST15 is widely reported in SouthEast Asia causing clonal outbreaks whereas it showed a different pattern in our setting driving smaller expansions but also as a lot of sporadic isolates occurring over time. This highlights the need for more data from sSA to understand the complex diversity of this organism in high-risk areas, as not only the ST composition, but also their patterns, differ between settings. The two key insights we gain from the population pattern is that the lineages continue to replace each other as the dominant clone over longer time periods, and that in our setting, this expansion is mainly driven by an increase in cases in neonatal patients. Most of the major sequence types are present at a steady, much lower proportion, before and after their respective clonal expansions, emphasizing the relevance of understanding what leads to the wave-like pattern of clonal expansions in *Kpn* populations.

This temporal variability also favours prioritisation of IPC strategies to prevent the introduction of new lineages. It may be that increased IPC efforts in response to ward outbreaks contribute to or are the single cause for the sudden stops of clonal expansions. We have little understanding of the ability of *Kpn* to survive on abiotic surfaces, which represents one of the key differences between HIC and LMIC settings, as the latter often use wooden furniture and bristled plastic tubing due to limited resources, which might lead to different *Kpn* lineages being successful. A similar question remains open regarding agents used for disinfection, as resource-limited settings often rely on basic soap agents and use unsafe water, whilst HIC routinely use industrially produced, quality controlled disinfectants.

Our data allow insight into AMR patterns in our setting. Ceftriaxone resistance, almost exclusively due to the ESBL-encoding *bla*_CTX-M-15_, is widespread. Whilst carbapenems are rarely available and resistance is already emerging (38,39), several treatment options are still viable, such as amikacin. Our analysis also emphasizes again that aminoglycoside resistance is notoriously hard to predict and might be impacted by the genetic background (for example efflux pumps) and not only acquired resistance genes, and should therefore at this stage not be ruled out based on genomic data but confirmed via phenotyping. Most isolates were phenotypically resistant to gentamicin; whether this is consequent on the predicted resistance genes, or whether other mechanisms drive this resistance is important to determine to improve the value of genotypic predictions for aminoglycosides. It will remain to be further investigated whether this is due to other interfering genes or low expression of the resistance gene and might be an indication of heteroresistance in the population which needs to be carefully considered(40). We furthermore observe a trend in re-emergence of chloramphenicol sensitivity, putatively in part driven by the spread of a non-functional CAT copy. Continuing to monitor re-emergence of susceptibility to agents no longer in use is of high relevance to allow timely re-introduction of drugs and can give us insights into the potential of rotating antimicrobial regiments over longer time spans to allow sensitivity to re-emerge.

There was little resistance predicted and observed to colistin, a last-line treatment option, in QECH *Kpn* isolates. Especially the lack of mobile resistance elements is encouraging, but also stresses the urgent need for better management of animal health as colistin is widely used, knowingly and as part of food products farmers are not necessarily aware of, in the increasing mid-size farming industry(41). Another exciting new development are initial results of combining fosfomycin and flomoxef(42); we observe only sporadic fosfomycin resistance mechanisms and very little spread of *ampC* genes, the only non-carbapenemases that would deactivate flomoxef, although we note that there are also counterindications for fosfomycin use in invasive infections.

As extensively resistant bacterial pathogens emerge and spread, there is an increasing urgency to develop vaccines to break the arms race between new antimicrobials and new resistance mechanisms. In this case there is interest in developing maternally administered *Klebsiella* vaccines to prevent neonatal sepsis. A target of high interest are the extracellular polysaccharides that can be typed based on their respective operons, which are assigned different K-types (capsular antigen) and O-types (the lipopolysaccharide O-antigen chain). The K-antigen shows a very high diversity; and whilst we can identify several dominant types in our study, the long timeframe of our study highlights the importance of considering the dynamic nature of K-antigens when considering vaccine design. Here, temporal fluctuations detected just in Blantyre strongly suggest that a K-type vaccine would become ineffective very rapidly given the frequent changes in dominant K-type, which might be driven by phage pressure in the environment and thus highly unpredictable. It is also unclear if successful vaccination against the major polysaccharide types would lead to a degree of cross protection against non-targeted types or whether lack of such cross protection would facilitate rapid emergence of infections caused by vaccine escape lineages.

In contrast, there are far fewer O-antigen types, making this a more attractive target. Recent efforts have compared several large datasets with O-and K-typing either determined by PCR or WGS(43); and identified that a conjugate vaccine targeting O1v1/2 (∼serotype O1)(31), O2v2 (∼serotype O2afg)(31) and O3b (∼serotype O3b)(31) has the potential to prevent the majority (>70%) of cases of *Kpn*-BSI. For the most at-risk population in our setting, the neonates, this would cover 87% of cases (338/388) assuming O1a and O1b were included into O1 and no other subtypes were discovered(30); importantly it would however not have prevented the ST340 outbreak in the neonatal unit previously described(15) (O4). It thus seems of relevance to consider including the currently low-abundance O-antigens as well, in particular given the interchangeability for O-types within the same STs. Reports from other highly understudied settings also report other main O-types in hospital isolates, e.g. O5 in a study in Pakistan(17,44) or a diversity of O-types in the Caribbean where this O-Ag vaccine would only cover 62% (162/260) (45). Selection pressure might not directly drive evolution given that *Kpn* is not human restricted but could likely select for strains with currently less abundant O-types to increase in prevalence. Further, there remains an open question of how accessible the O-antigens are to antibodies, which a recent study calls into question(46); we currently have very limited understanding what drives the extent of capsule expression per cell in non-hypermucoid isolates and how different stages of the infection impact this. It is known from *in vitro* studies that capsule expression even in the same experimental setting can differ widely within clonal lineages, which also impacts the bacterial ability to escape the complement system(47).

Our data, and the recent identification of a novel form of the long-studied O1 antigen type(30,48–50), highlights that we have very little understanding of the relevance of the O-and K-types on the success of *Kpn* lineages, whether any additional genes on the chromosomal backbone can further modify the structures and lead to different serotypes with the same O-locus, as well as whether any O-/K-combinations are more or less favourable. Closing such knowledge gaps will be central to design of vaccines as we also have little understanding for the propensity of sequence type combinations with specific vaccine target serotypes such as capsule or O-antigen. A more solid understanding of O-/K-type variations, both regarding their biochemistry and whether they contribute to epidemiological success, is crucial given the high interest to use these as targets for maternally administered vaccines against neonatal sepsis.

This highlights that we have very little understanding of the relevance of the O-and K-types on the success of Kpn lineages, whether any additional genes on the chromosomal backbone can further modify the structures and lead to different serotypes with the same o-locus, as well as whether any O-/K-combinations are more or less favourable. A more solid understanding of O-/K-type variations, both regarding their biochemistry and whether they contribute to epidemiological success, is crucial given the high interest to use these as targets for maternally administered vaccines against neonatal sepsis.

## Conclusions

In summary, our study provides a unique insight into long-term patterns of *Kpn* hospital isolates from a low-income setting, spanning across a time period where the number of MDR and ESBL producing infections increased rapidly across the globe. We show that whilst STs known as high-risk in other settings are common, these do not always show similar success in different settings; and we observe a significant numbers of isolates from otherwise rarely observed STs, causing hospital infections. This again emphasizes the importance of including the relevant settings into studies aiming to identify treatment or vaccine targets as we currently have poor understanding of what makes an ST a high-risk lineage in each setting at a given time. Our data also emphasizes the urgent need to consider temporal variation when considering treatment or vaccine targets, as some of the main vaccine targets show strong temporal fluctuations; the most prevalent type now circulating is unlikely to cause the next outbreak. The close relatedness of isolates from the same ward in the same time frame strongly indicate that the majority of infections are hospital-acquired, so whilst vaccine development and alternative treatment options are highly relevant; at this stage, focused efforts to improve IPC are key to reducing *Kpn* disease burden now(21).

## Methods

### Isolate collection

QECH is a government run tertiary referral centre for the southern region of Malawi, which provides free care at the point of delivery. It serves a population of 800,000 for the area of urban Blantyre (Malawi census 2018). There are adult and paediatric A&Es, medical and surgical wards. There is a tertiary neonatal unit on site and a separate paediatric surgical hospital which opened in 2017 (Figure 2C; PICU, Paediatric HDU, Paediatric surgical ward).

### Microbiological sampling and processing

Routine, (ISO15189 accredited since 2019) diagnostic blood culture services have been provided to the medical and paediatric wards by the Malawi-Liverpool-Wellcome Programme since 2000. For adults 7-10mL of blood were collected from all patients admitted to the hospital with fever (axillary temperature >37.5°C) or clinical suspicion of sepsis, severe sepsis, or septic shock. Sepsis, severe sepsis, or septic shock were suspected in patients with tachycardia (≥90 beats per minute), hypotension (systolic blood pressure <90 mm Hg), tachypnoea (respiratory rate >20 per minute), or delirium. 3–10 mL of blood was taken from children with non-focal febrile illness who tested negative for malaria, who were severely ill with suspected sepsis, or who failed initial malaria treatment and remained febrile. For adults and children with clinical suspicion of meningitis (temperature >37.5°C with seizures, headache, abnormal behaviour or meningism) a lumbar puncture was also performed and the sample sent for CSF analysis. For neonates with clinical signs of sepsis or meningitis (temperature >37.5°C, or other signs of clinical deterioration such as reduced activity, increasing work of breathing or respiratory rate >60, heart rate >180 or <100, and unexplained seizures) a blood culture and CSF sample was collected. Afebrile patients were unlikely to have had blood or CSF cultures unless the clinical suspicion for sepsis or meningitis was high.

For blood cultures, samples were collected using aseptic methods and inoculated into a single aerobic bottle (BacT/Alert, bioMérieux, Marcy-L’Etoile, France). These were incubated using the automated BacT/Alert system (bioMérieux, France) since 2000. Before which, they were cultured manually. Samples that flagged positive were Gram stained and Gram-negative bacilli are identified by Analytical Profile Index (Biomérieux). CSF samples were processed for cell count and biochemistry before manual culture. Antimicrobial susceptibility testing was determined by the disc diffusion method (Oxoid, United Kingdom) using AST breakpoints that were current at the time, initially following the relevant version of the British Society of Antimicrobial Chemotherapy’s guidelines until 2018, after which time EUCAST methods and breakpoints were introduced. Organisms which showed reduced inhibition when exposed to Ceftriaxone 30mg discs, Ceftriaxone 5mg discs, or Cefpodoxime 10mg discs were classified as resistant to Ceftriaxone. Intermediate isolates were classified as resistant to ceftriaxone. Details for the identification of other organisms are described elsewhere(3). *Klebsiella pneumoniae* isolates were stored at -80°C on microbank beads.

### Whole genome sequencing and QC

All *Klebsiella pneumoniae* isolates that could be recovered from the start of the MLW archive to April 2020 were thawed and incubated on MaConkey’s media for 18-24 hours at 37°C. Plates with pure growth then had a single colony pick taken and inoculated into 15 ml of buffered peptone water for 18-24 hours at 37°C. These samples were then centrifuged and the supernatant was discarded. The pellet was then resuspended in buffer and the DNA was extracted using the QIAsymphony machine and QIAsymphony DSP kit with onboard lysis, according to the manufacturer’s instructions. Quality control was done using Qubit and samples with a DNA volume of less than 200ng were repeated. DNA was sequenced at the Wellcome Sanger Institute on the Illumina HiSeq X10 instrument (Illumina Inc., United States) to produce 150 bp paired end reads.

Eleven out of 1313 samples failed the Sanger-internal sequencing threshold for minimum coverage for assemblies and were thus removed from further analyses. Quality control on all remaining isolates was performed using species confirmation via Kraken v0.10.6(51) to identify contaminants (incorrect species). Any species assignment other than *Klebsiella pneumoniae*, *Klebsiella variicola*, unclassified and synthetic construct were considered contaminants; the latter two categories frequently derive from plasmid sequences in our experience. All isolates with higher than 10% of other species were removed as contaminated (Fig S2A). Similarly, isolates with a ratio of heterozygous SNPs over 5% were removed as likely contaminated by a related strain (Fig S2B). Further quality control used the assemblies, where all isolates resulting in more than 1000 contigs were removed as likely contaminated by other species or strains, or of very low quality (Fig S2C). *De novo* assembly of the remaining 1156 genome sequences was performed using SPAdes v3.14.0(52) incorporated into the bacterial genome assembly pipeline described previously(53). Annotation of all assemblies was performed using prokka(54) v1.14.5.

### Genome analyses and phylogenetics

For comparative analyses, isolates from recent studies at QECH were included, as they are part of the bloodstream collection(14–16). Details of all sequences used in the analyses are given in supplementary table 1. Resistance gene prediction and plasmid replicon types were performed using ariba(55) (v2.14.6) and the srst2-argannot(56,57), CARD(58) and plasmidfinder(59) databases (download date 14.02.2022). To assess aminoglycoside resistance; ANT3 is by ariba as AacAad_AGly, AadA4_5_AGly, AadA_AGly, AadA_AGly_2, AphA6_AGly; AAC(3)-II is grouped by ariba as Aac3_IIa_AGly; ANT(2”)-I is grouped by ariba as AadB_AGly and AAC(6’)-I is grouped by ariba as AacAad_AGly_3.

Prediction of sequence types, (sub)species and *Klebsiella*-specific virulence factors were performed using kleborate v2.1.0(60) and the –r setting; the results are given for all isolates in Table S1. O-and K-types were predicted with kaptive(31) (v2.0.6) which incorporates O-loci as well as O-serotype prediction(29,31,61); and we added an additional option of *wbbZ* as extra gene to distinguish between O1a (O1/O2 + *wbbY*, no *wbbZ*) and O1b (O1/O2 + *wbbY* + *wbbZ*) as described recently(30), adding *wbbZ* as implemented in kaptive versions before v2(61) and expanding the .logic file to distinguish O1a/b.

Core genome SNPs for *K. pneumoniae* subsp. *pneumoniae* isolates were determined using snippy(62) (v4.6.0; https://github.com/tseemann/snippy) with the *Klebsiella pneumoniae* subsp. *pneumoniae* NTUH-K2044 genome (AP006725.1)(63) as reference. Recombinant regions were removed using gubbins(64) (v3.2.1) and the FastTree(65) option (‘-tree-builder fasttree’), given the very high number of samples, as implemented in gubbins; and only isolates from sequence types with more than one isolate in our dataset were included in the analysis to further reduce compute time. The same strategy was used for *K. quasipneumoniae* and *K. variicola* using strains ATCC 700603 (CP029597.1)(66) and At-22 (CP001891.1)(67) as reference, respectively, where all isolates were included and default options for gubbins were used (RaxML for tree generation, given the much smaller sample numbers). A snp-only alignment was generated from the recombination-free gubbins output using snp-sites(68) v2.5.1 with the -c option (selecting for ACGT-only) for *K. quasipneumoniae* and *K. variicola*; for *K. pneumoniae* subsp. *pneumoniae*, given the high variation of isolates, the -c option was too stringent (resulting in a <200 bp alignment) and we thus used snp-sites to isolate all snps but without selecting for ACGT-only sites.

The snp-only alignments were used as input for iqtree(69) (v1.6.12), the number of constant sites was determined with snp-sites -C function from the original full alignment and provided to the ‘-fconst x,x,x,x’ iqtree function. The model was determined with the integrated ModelFinder(70) function ‘-m TEST’ resulting in GTR+F+I+G4, GTR+F+I+G4, TVM+F+I+G4, for the alignments from *K. pneumoniae*, *K. quasipneumoniae* and *K. variicola*, resp. We set ‘- keep-ident’ to keep identical data and ‘-bb 1000’ for 1000 ultrafast bootstrap replicates. The resulting trees were then adjusted to SNP-length branch lengths using the python implementation of pyjar(71) (https://github.com/simonrharris/pyjar) as input for rPinecone(19). Closely related clusters taking their phylogenetic distance into account were determined with 5 SNPs as cut-off for minor and 10 as cut-off for major clusters. These were visualised with iTol(72) (v6.8) with visualisation files generated as implemented in rPinecone(19). For ST35, ST39 and ST14/15 (combining ST14 and ST15 isolates in one analysis) we performed ST-specific mappings of all respective isolates against references from these sequence types to gain higher resolution (CP041353.2(73), LR991401.1(74) and CP008929.1(75), resp.).

Pan-genome reconstruction, generation of a core gene alignment, and identification of neighbouring gene regions were derived via panaroo(76) (v1.3.3) using the ‘–clean-mode strict’ setting. The phylogenetic tree based on the core gene alignment, with informative sites extracted with snp-sites as above, was calculated using iqtree(69) (v1.6.12) and the internal ModelFinder(70) function (GTR+F+ASC+G4) as above. All other metadata summary analyses and visualisation were generated using the ggplot(77) and ggtree(78) packages in R(79) unless indicated otherwise; some colour schemes are derived from or inspired by the MetBrewer (https://github.com/BlakeRMills/MetBrewer) and NationalParks (https://github.com/katiejolly/nationalparkcolors) R libraries. Composite figures were generated using the patchwork package (https://github.com/thomasp85/patchwork). An RMarkdown file to generate the analysis plots and the underlying tree and data files are freely available online (https://github.com/EvitaH/QECHospitalKlebs; doi: 10.5281/zenodo.8421658).

## Data availability

All sequence data is available on SRA/ENA under BioProject PRJEB42462; specific accessions and all additional data is provided in the supporting tables S1-S5. All data and code used to create the analyses and is available as R package QECHospitalKlebs v0.0.9 (https://github.com/EvitaH/QECHospitalKlebs; doi: 10.5281/zenodo.8421658).

## Data Availability

All data produced in the present work are contained in the manuscript and available in online repositories as described in the manuscript.

https://www.ebi.ac.uk/ena/browser/view/PRJEB42462

## Acknowledgments

We would like to thank the Sanger Institute Pathogen Genomics group for expert computational support. We acknowledge and thank all the clinical staff and patients at QECH over the years for their contribution; and the MLW core microbiology team for expert support in storage and handling of the isolates.

## Ethics statement

Ethical approval for this study was granted by the University of Malawi College of Medicine Research Ethics Committee (COMREC) (P.11/18/2541) and isolates were shipped under a Nagoya Protocol compliant Access and Benefit Sharing agreement between the Government of Malawi and the Wellcome Sanger Institute.

## Author contributions

Conceptualization: *EH, NAF;* Methodology: *EH, NAF;* Software: *EH, JML;* Validation: *EH, OP, AZ;* Formal analysis: *EH, OP;* Investigation: *EH, OP, AZ, PM, PS, ET, FG, RL, SL, JC, JML;* Resources: *SB, PS, ET, KK, NRT, NAF;* Data curation: *EH, OP, PM, RL, JC, JML, AZ;* Writing—original draft: *EH, OP, NAF;* Writing—review & editing: *all authors;* Visualization: *EH, OP, JML, NAF;* Supervision: *EH, CM, SL, KK, NAF;* Project administration: *EH, NAF;* Funding acquisition: *EH, NRT, NAF*

