## Supplementary Figures for "Longitudinal analysis within one hospital in sub-Saharan Africa over 20 years reveals repeated replacements of dominant clones of *Klebsiella pneumoniae* and stresses the importance to include temporal patterns for vaccine design considerations"

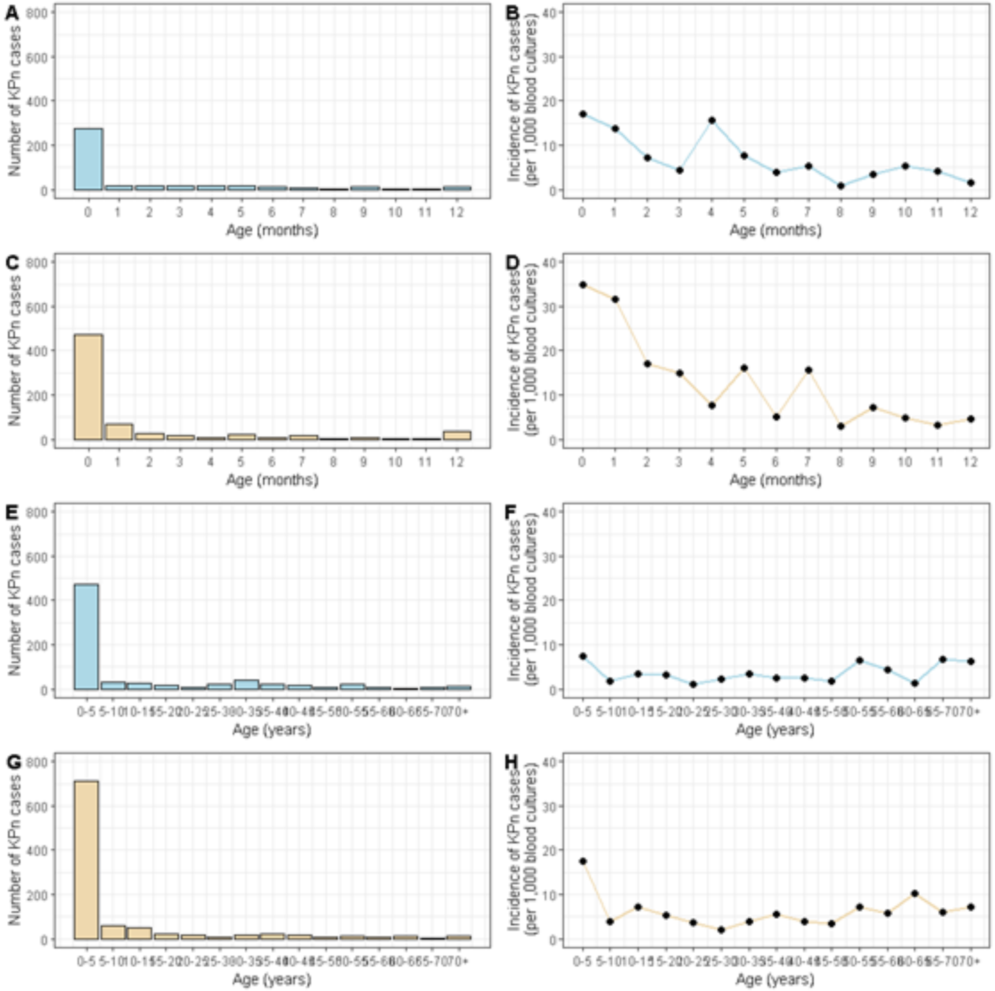

A
B
C
D
E
F
G
H
Number of KPn cases
Incidence of KPn cases (per 1,000 blood cultures)
Age (months)
Age (years)

**Fig S1. Age specific *Kpn* crude frequency and blood culture positivity rate.** (**A**) Number of *Kpn* cases up to age 12 months (1998-2015). (**B**) Blood culture positivity rate (per 1,000 blood cultures) of *Kpn* cases up to age 12 months (1998-2015). (**C**) Number of *Kpn* cases up to age 12 months (2016-2021). (**D**) Blood culture positivity rate (per 1,000 blood cultures) of *Kpn* cases up to age 12 months (2016-2021). (**E**) Number of *Kpn* cases up to 100 years (1998-2015). (**F**) Blood culture positivity rate (per 1,000 blood cultures) of *Kpn* cases up to 100 years (1998-2015). (**G**) Number of *Kpn* cases up to 100 years (2016-2021). (**H**) Blood culture positivity rate (per 1,000 blood cultures) of *Kpn* cases up to 100 years (2016-2021).

Figure S2

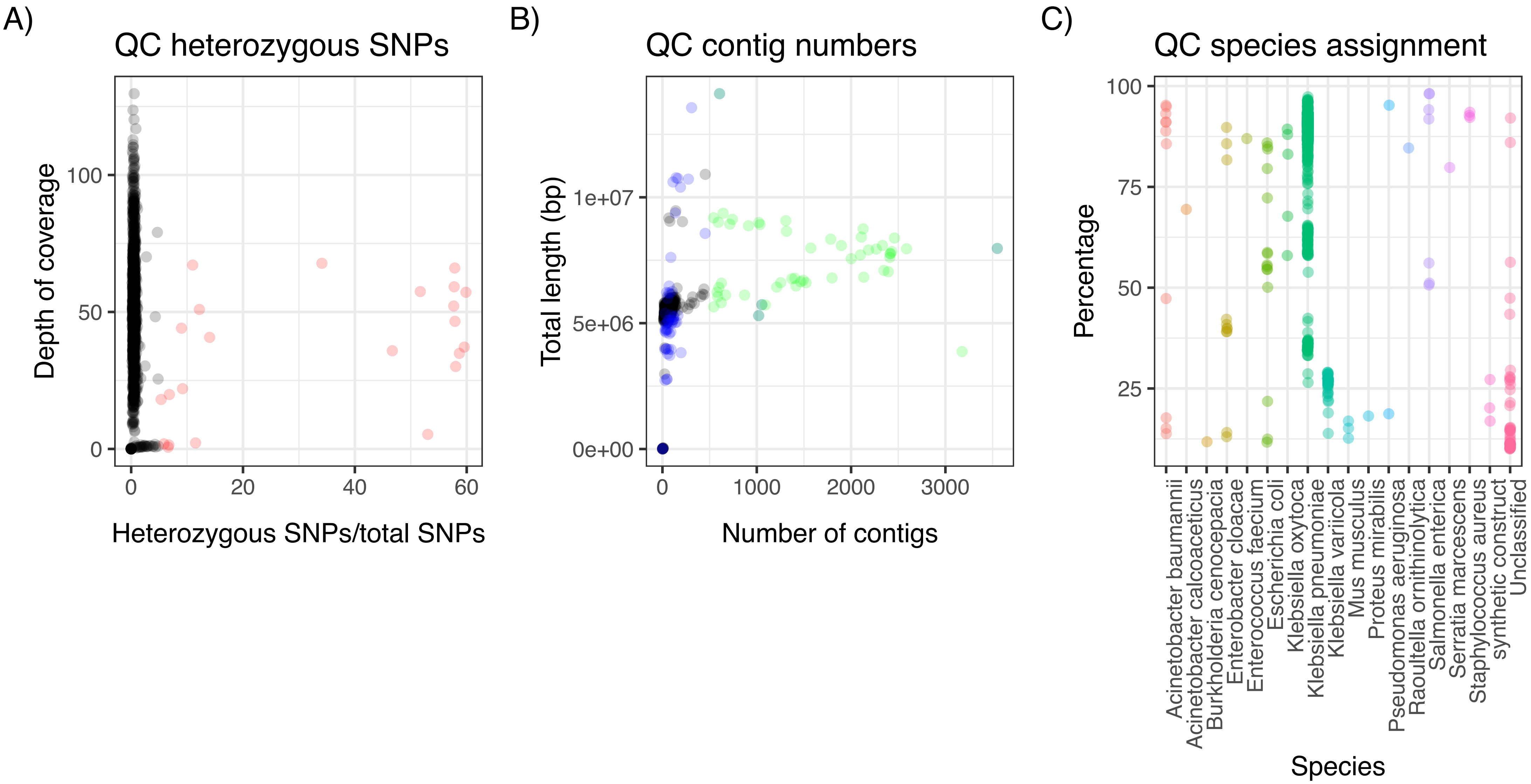
A)
QC heterozygous SNPs
Depth of coverage
Heterozygous SNPs/total SNPs
0
50
100
0
20
40
60
B)
QC contig numbers
Total length (bp)
Number of contigs
0e+00
5e+06
1e+07
0
1000
2000
3000
C)
QC species assignment
Percentage
Species
25
50
75
100
Acinetobacter baumannii
Acinetobacter calcoaceticus
Burkholderia cenocepacia
Enterobacter cloacae
Enterococcus faecium
Escherichia coli
Klebsiella oxytoca
Klebsiella pneumoniae
Klebsiella variicola
Mus musculus
Proteus mirabilis
Pseudomonas aeruginosa
Raoultella ornithinolytica
Salmonella enterica
Serratia marcescens
Staphylococcus aureus
synthetic construct
Unclassified

**Fig S2. Quality control of sequencing data.** The species assignment of the sequencing reads according to kraken are shown in panel (**A**), where all isolates with more than 10% of a different species were removed. (**B**) The plot shows the distribution of homozygous vs heterozygous SNPs, isolates represented by red points (>5% heterozygous SNPs) were removed as likely consisting of two related strains. (**C**) Number of contigs vs total assembly length is shown; with isolates that passed QC in black, isolates that failed the species composition QC in blue, and isolates with >500 contigs in green, which were also removed.

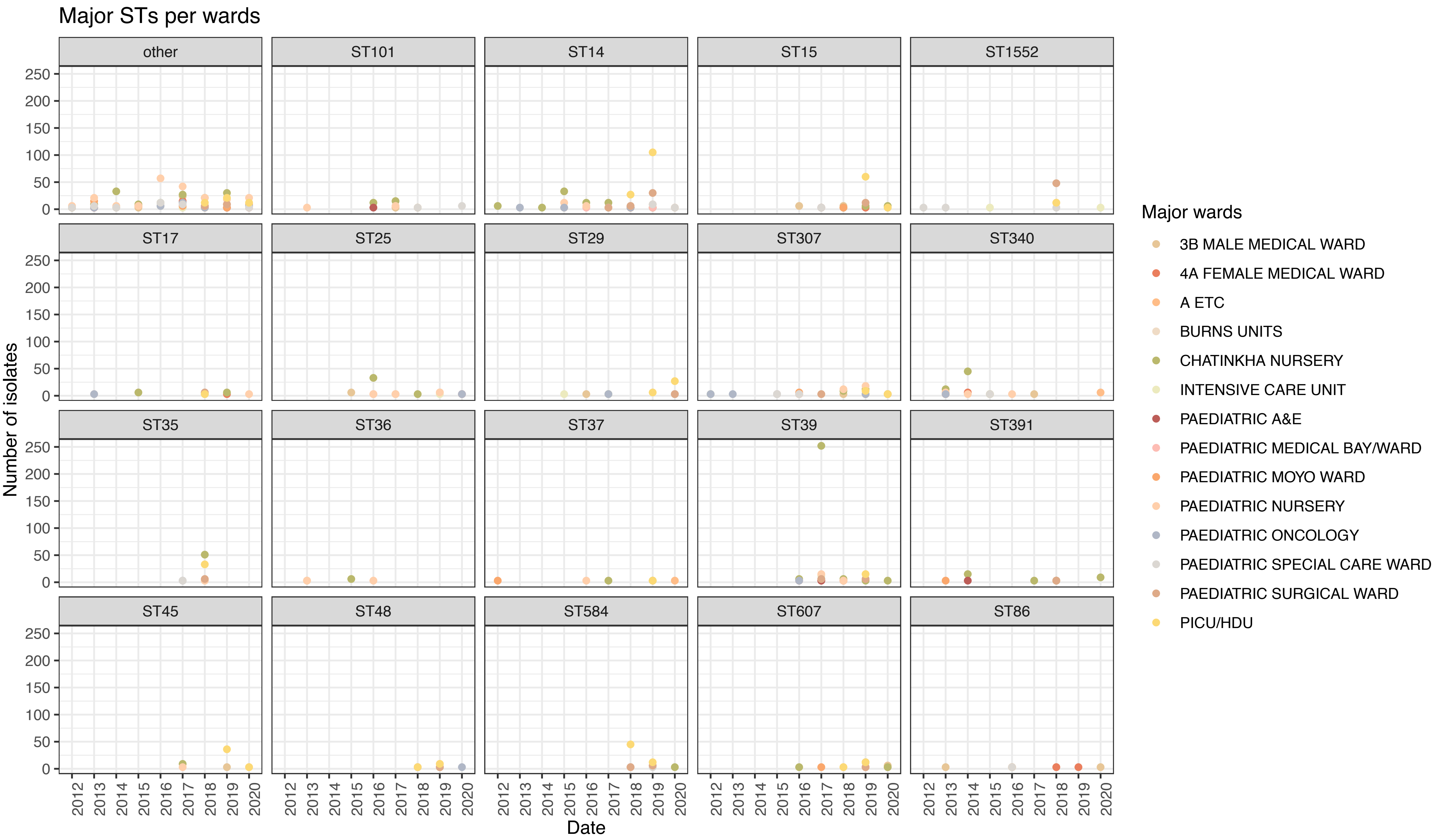

Major STs per wards
other
ST101
ST14
ST15
ST1552
ST17
ST25
ST29
ST307
ST340
ST35
ST36
ST37
ST39
ST391
ST45
ST48
ST584
ST607
ST86
Number of isolates
0
50
100
150
200
250
2012
2013
2014
2015
2016
2017
2018
2019
2020
Date
Major wards
3B MALE MEDICAL WARD
4A FEMALE MEDICAL WARD
A ETC
BURNS UNITS
CHATINKHA NURSERY
INTENSIVE CARE UNIT
PAEDIATRIC A&E
PAEDIATRIC MEDICAL BAY/WARD
PAEDIATRIC MOYO WARD
PAEDIATRIC NURSERY
PAEDIATRIC ONCOLOGY
PAEDIATRIC SPECIAL CARE WARD
PAEDIATRIC SURGICAL WARD
PICU/HDU

**Fig S3. Main ST isolates per wards over time.** Each panel shows the number of isolates for the relevant ST as in the legend. The number of isolates per year are stratified by the major wards as in the figure legend.

Figure S4

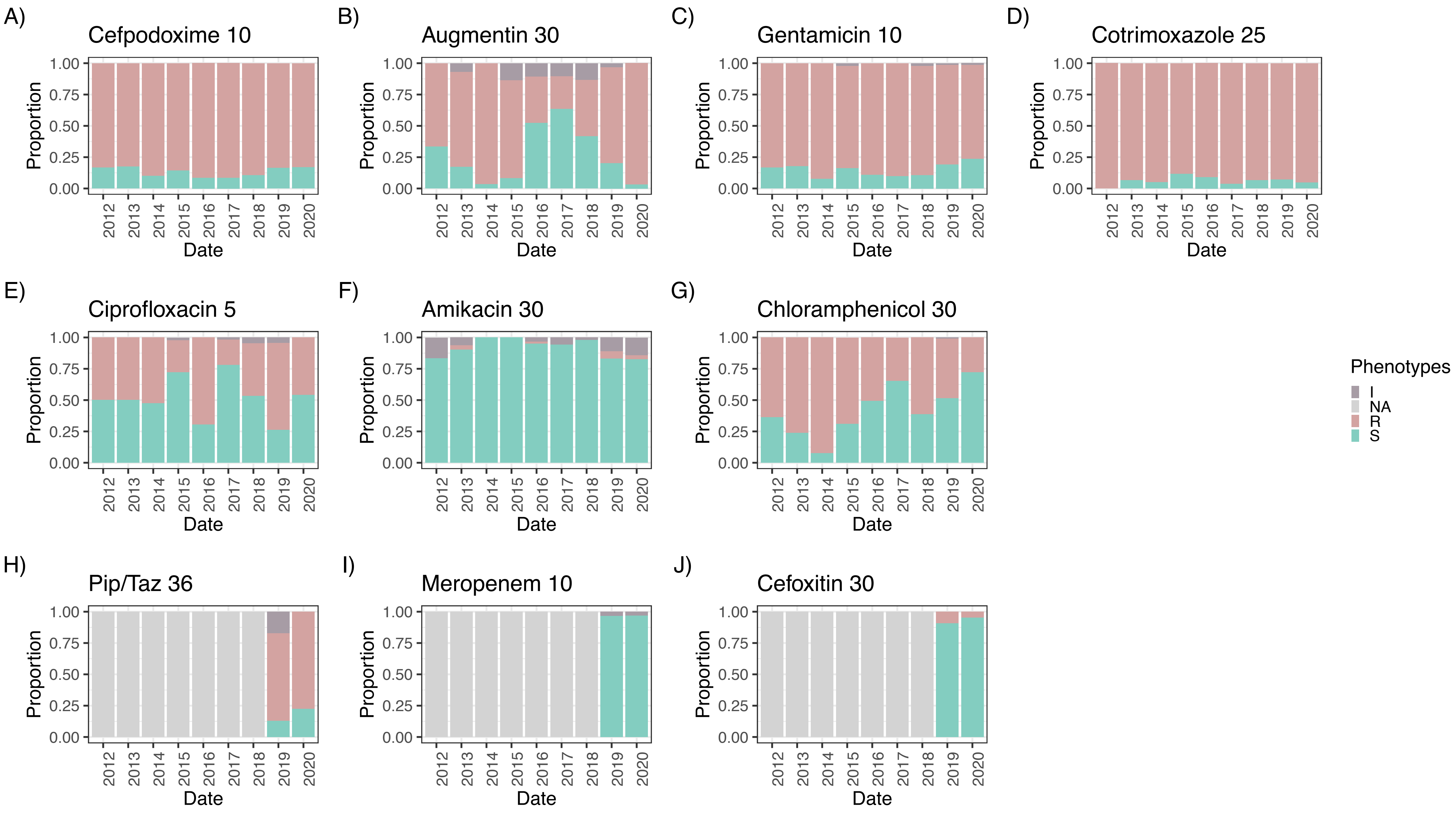
A)
Cefpodoxime 10
B)
Augmentin 30
C)
Gentamicin 10
D)
Cotrimoxazole 25
E)
Ciprofloxacin 5
F)
Amikacin 30
G)
Chloramphenicol 30
H)
Pip/Taz 36
I)
Meropenem 10
J)
Cefoxitin 30
Proportion
1.00
0.75
0.50
0.25
0.00
2012
2013
2014
2015
2016
2017
2018
2019
2020
Date
Phenotypes
I
NA
R
S

**Fig S4. Phenotypic resistance profiles of all sequenced isolates**. This highlights very high resistance against widely used first-line treatments like (**A**) cefpodoxime (ceftriaxone; 3GC), (**B**) augmentin (BLBLI combination), (**C**) gentamicin (AGly), (**D**) cotrimoxazole (TMT) but still a large proportion of sensitive isolates against less commonly used antimicrobials; (**E**) ciprofloxacin (Fq), (**F**) amikacin (AGly), and a pattern of increasing sensitivity against (**G**) chloramphenicol. The currently only available last-line treatments are (**H**) piperazillin-tazobactam (BLBLI combination), which show very high resistance levels; and (**I**) meropenem (carbapenem), the currently only viable alternative tested. (**J**) Cefoxitin also shows a lot of sensitive isolates; this antimicrobial is routinely used in diagnostic laboratories world-wide to test for *ampC* production as it was rapidly replaced with third-generation cephalosporins after its initial characterisation. The resistance profile is however of high interest as clinical trials have been started recently to assess it as a potential treatment alternative.

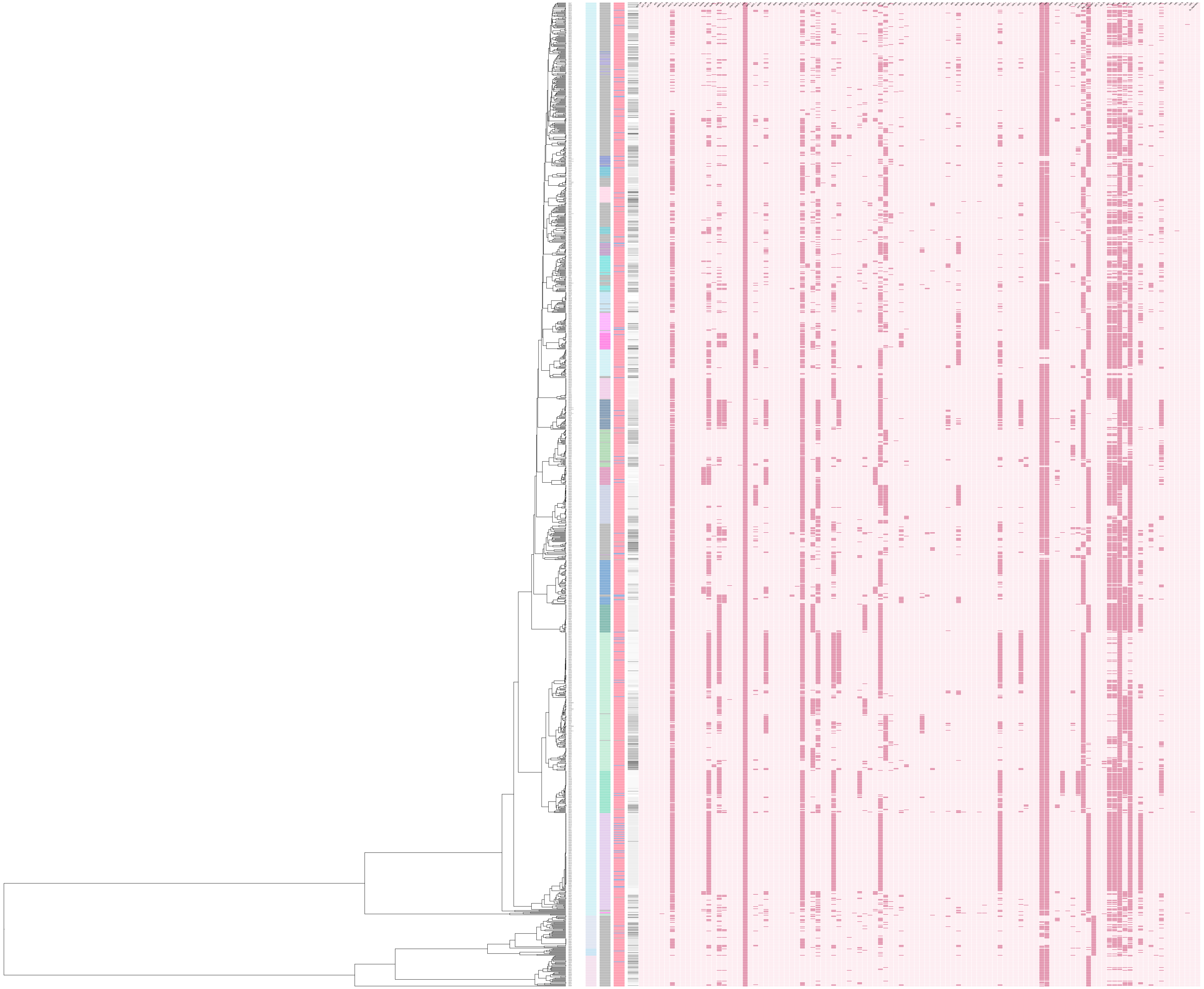

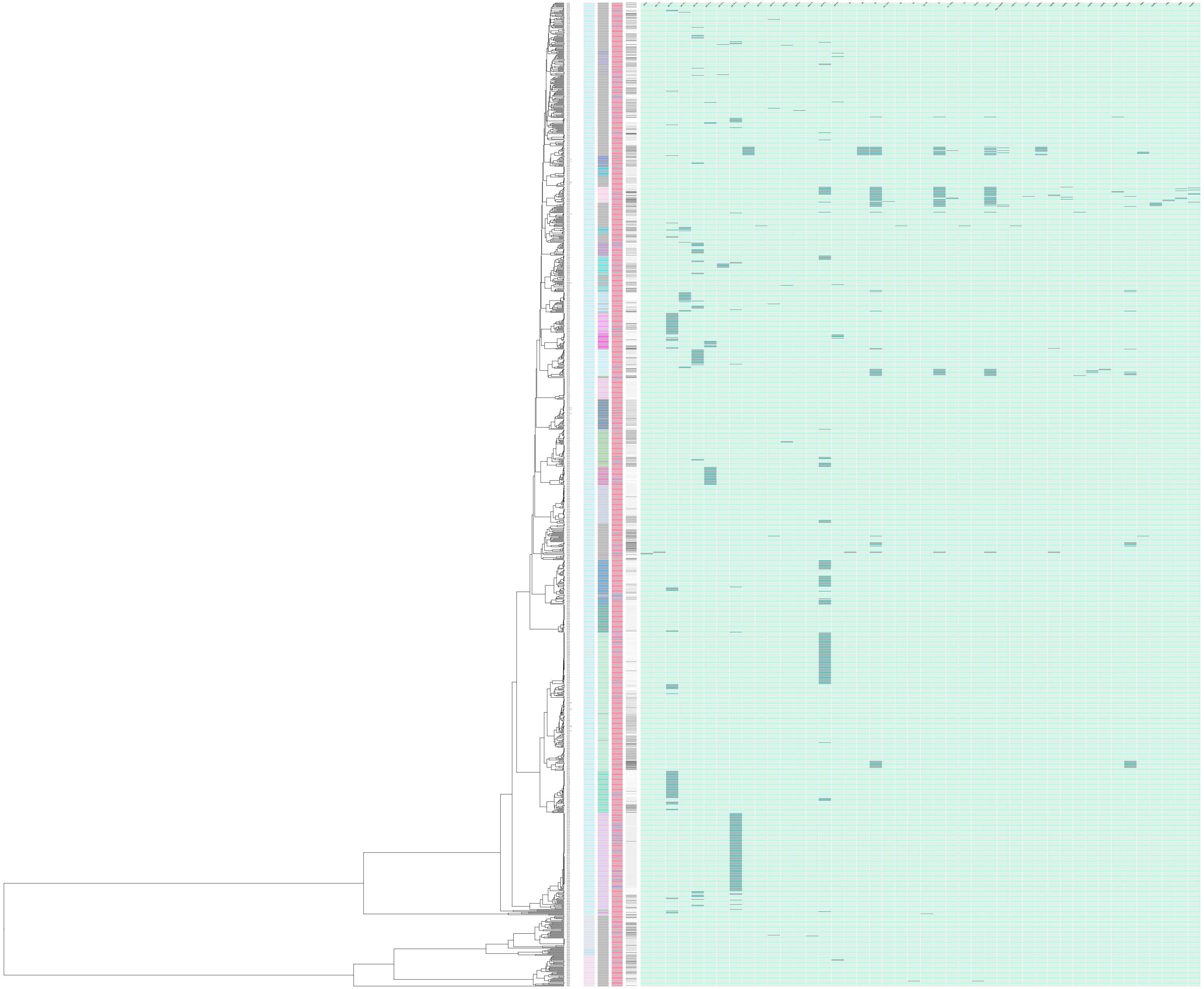

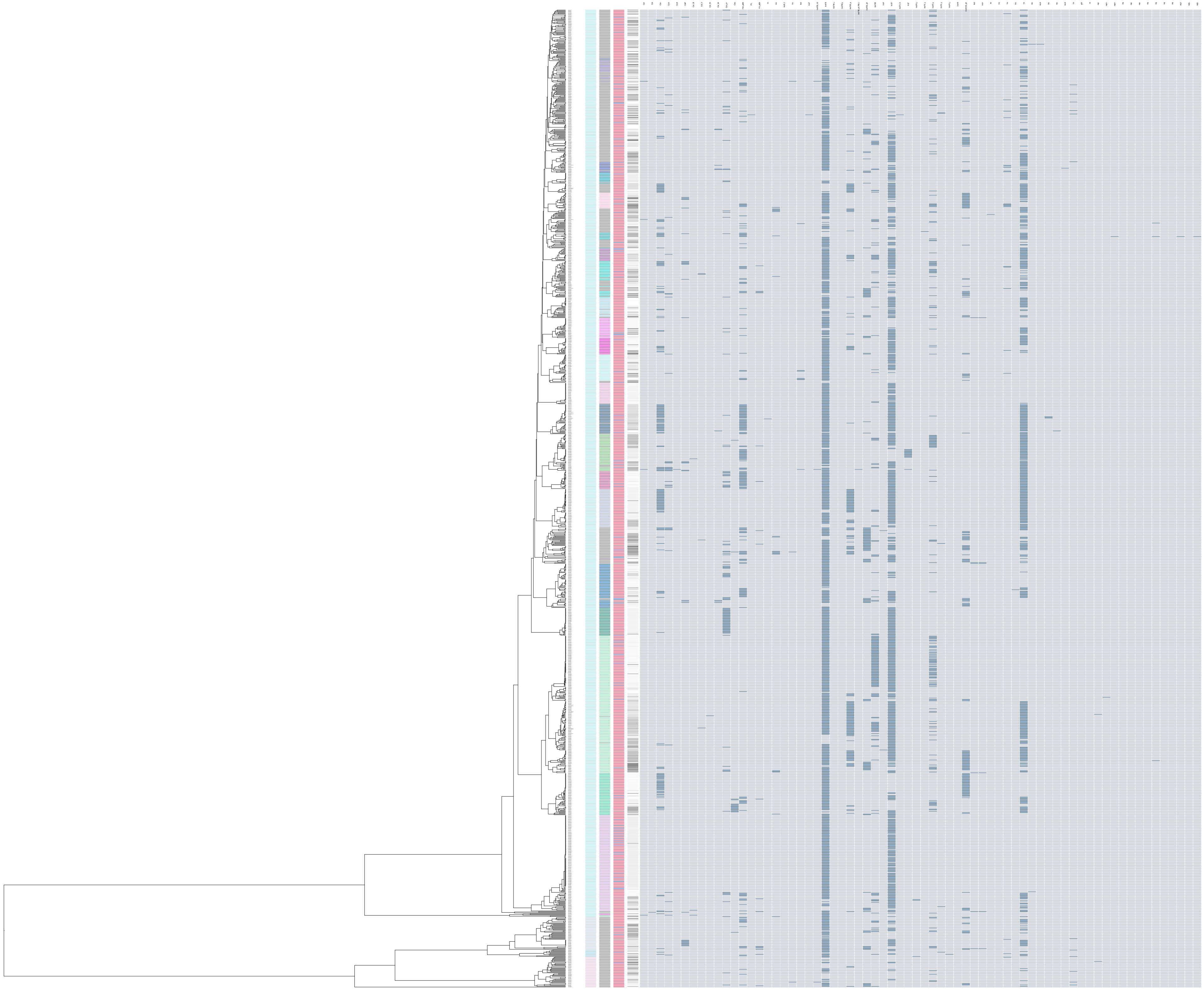

**Fig S5. Predicted resistance and virulence genes and plasmid replicons.** The guidance tree was calculated with iqtree based on the core gene alignment derived from panaroo. The metadata columns displayed are *Kpn* species, main ST, sample type and year of isolation; as well as heatmaps showing the presence (dark shade) or absence (light shade) of (**A**) resistance genes, (**B**) virulence genes and (**C**) plasmid replicons.

Figure S6

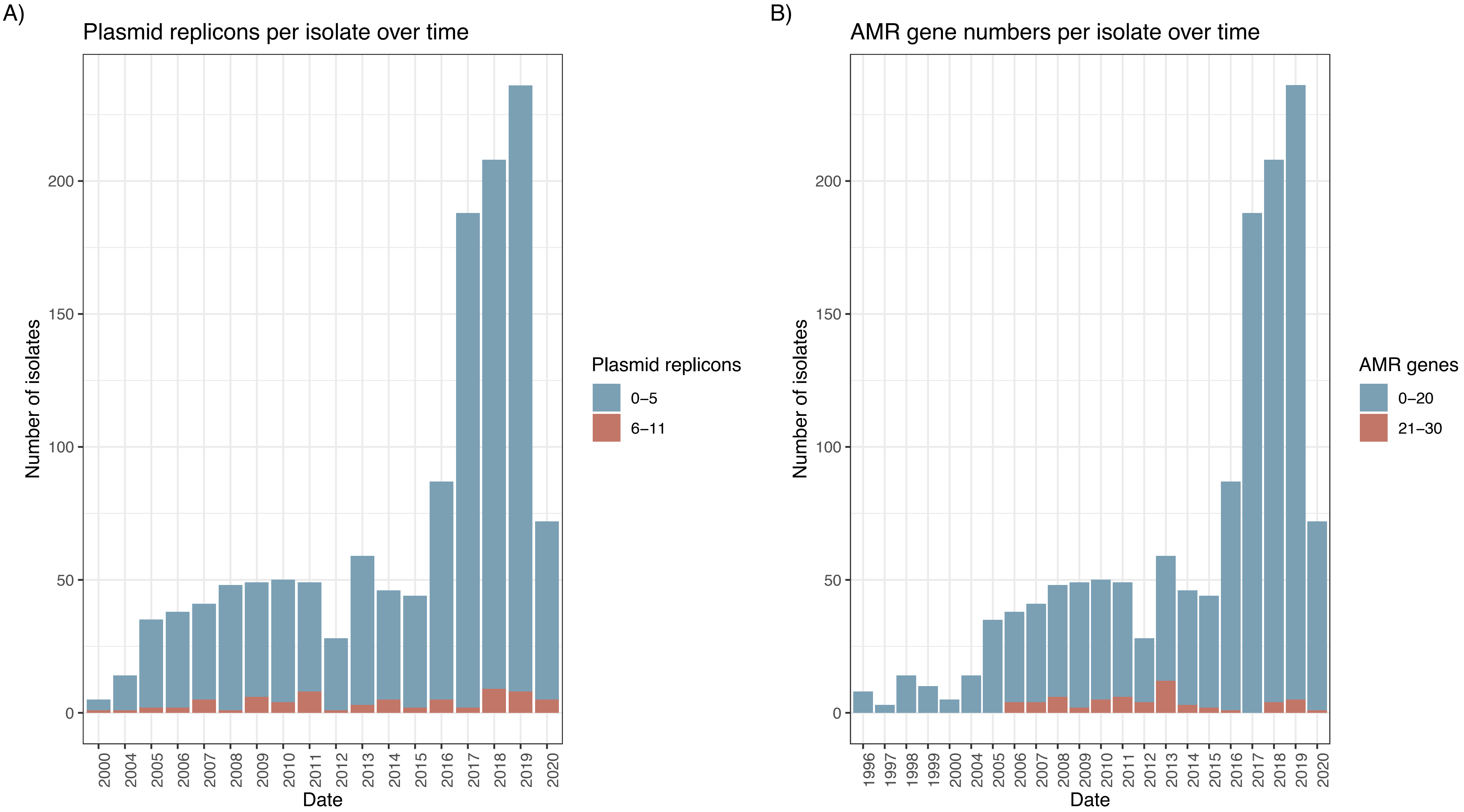
A)
Plasmid replicons per isolate over time
Number of isolates
0
50
100
150
200
2000
2004
2005
2006
2007
2008
2009
2010
2011
2012
2013
2014
2015
2016
2017
2018
2019
2020
Date
Plasmid replicons
0−5
6−11
B)
AMR gene numbers per isolate over time
Number of isolates
0
50
100
150
200
1996
1997
1998
1999
2000
2004
2005
2006
2007
2008
2009
2010
2011
2012
2013
2014
2015
2016
2017
2018
2019
2020
Date
AMR genes
0−20
21−30

**Fig S6. Plasmid replicons and resistance gene numbers.** (**A**) *Kpn* isolates over time showing the number of plasmid replicons encoded, and (**B**) the number of isolates for numbers of resistance genes present over time.

Figure S7

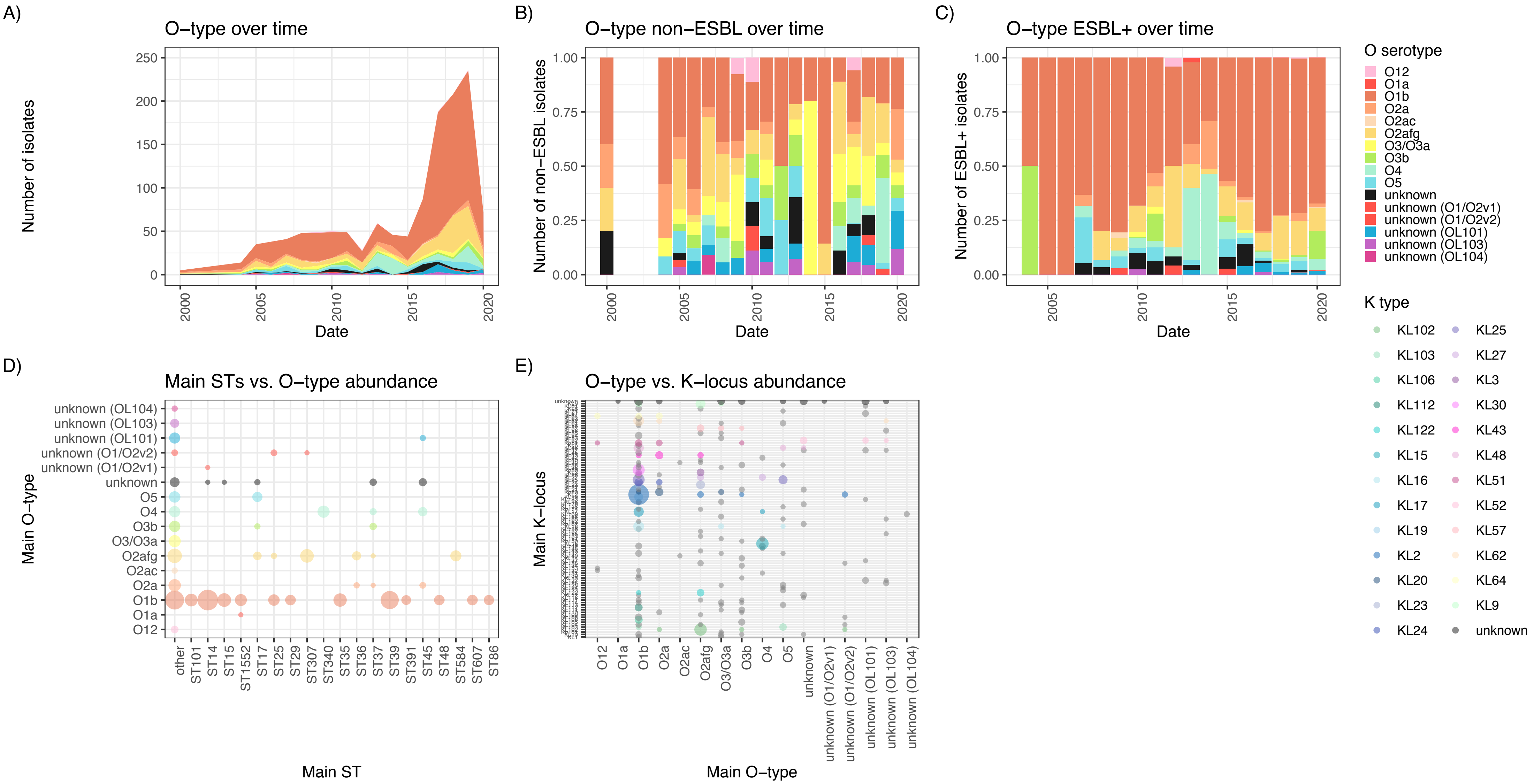
A)
O−type over time
Number of isolates
0
50
100
150
200
250
2000
2005
2010
2015
2020
Date
B)
O−type non−ESBL over time
Number of non−ESBL isolates
0.00
0.25
0.50
0.75
1.00
2000
2005
2010
2015
2020
Date
C)
O−type ESBL+ over time
Number of ESBL+ isolates
0.00
0.25
0.50
0.75
1.00
2005
2010
2015
2020
Date
O serotype
O12
O1a
O1b
O2a
O2ac
O2afg
O3/O3a
O3b
O4
O5
unknown
unknown (O1/O2v1)
unknown (O1/O2v2)
unknown (OL101)
unknown (OL103)
unknown (OL104)
K type
KL102
KL103
KL106
KL112
KL122
KL15
KL16
KL17
KL19
KL2
KL20
KL23
KL24
KL25
KL27
KL3
KL30
KL43
KL48
KL51
KL52
KL57
KL62
KL64
KL9
unknown
D)
Main STs vs. O−type abundance
Main O−type
unknown (OL104)
unknown (OL103)
unknown (OL101)
unknown (O1/O2v2)
unknown (O1/O2v1)
unknown
O5
O4
O3b
O3/O3a
O2afg
O2ac
O2a
O1b
O1a
O12
other
ST101
ST14
ST15
ST1552
ST17
ST25
ST29
ST307
ST340
ST35
ST36
ST37
ST39
ST391
ST45
ST48
ST584
ST607
ST86
Main ST
E)
O−type vs. K−locus abundance
Main K−locus
O12
O1a
O1b
O2a
O2ac
O2afg
O3/O3a
O3b
O4
O5
unknown
unknown (O1/O2v1)
unknown (O1/O2v2)
unknown (OL101)
unknown (OL103)
unknown (OL104)
Main O−type

**Fig S7. O-Ag serotypes over time and resistance profiles.** The distribution of predicted O-Ag serotypes based on their genomic loci(24,27) over time for (**A**) all isolates, (**B**) non-ESBL and (**C**) ESBL-encoding isolates, and their distribution across (**D**) main STs and (**E**) K-locus types.

Figure S8

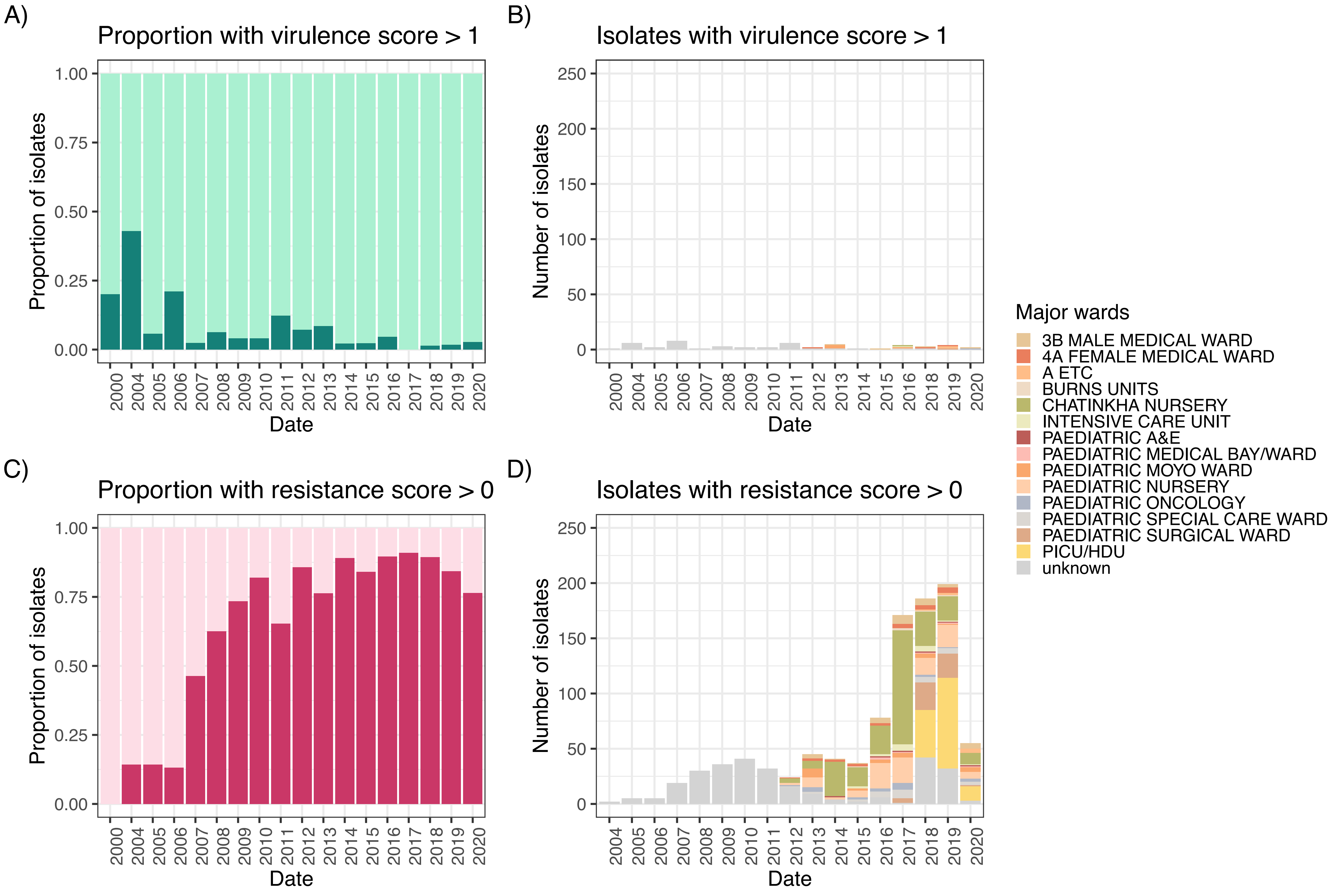
A)
Proportion with virulence score > 1
Proportion of isolates
1.00
0.75
0.50
0.25
0.00
2000
2004
2005
2006
2007
2008
2009
2010
2011
2012
2013
2014
2015
2016
2017
2018
2019
2020
Date
B)
Isolates with virulence score > 1
Number of isolates
250
200
150
100
50
0
Date
C)
Proportion with resistance score > 0
Proportion of isolates
Date
D)
Isolates with resistance score > 0
Number of isolates
Date
Major wards
3B MALE MEDICAL WARD
4A FEMALE MEDICAL WARD
A ETC
BURNS UNITS
CHATINKHA NURSERY
INTENSIVE CARE UNIT
PAEDIATRIC A&E
PAEDIATRIC MEDICAL BAY/WARD
PAEDIATRIC MOYO WARD
PAEDIATRIC NURSERY
PAEDIATRIC ONCOLOGY
PAEDIATRIC SPECIAL CARE WARD
PAEDIATRIC SURGICAL WARD
PICU/HDU
unknown

**Fig S8. Proportions of virulent isolates and resistant isolates over time have markedly different pattern.** (**A**) The number of isolates with a kleborate-predicted virulence score > 1 (1 usually indicates yersiniabactin which is chromosomally fixed in several STs) which remains very low and stable over time and found sporadically across various wards, whereas (**B**) isolates with resistance scores 1 or over (indicating ESBL or, in the case of score of 2, carbapenemase present) rapidly increase over time and biased towards several wards. This is reflected in the proportions; whilst isolates encoding several virulence factors remain at very low proportion over time (**C**), we can see the rapid increase of resistant isolates which remains at a very high level since the initial increase (**D**).

**Table S1:** Accessions, metadata and kleborate predictions per strain.
**Table S2:** Antimicrobial gene predictions using ariba.
**Table S3:** Plasmid replicon predictions using ariba.
**Table S4:** Phenotypic resistance profiles.
**Table S5**: O- and K-antigen predictions using updated definitions in kaptive.
